# Evaluating Large Language Models for Assessment of Psychosis Risk

**DOI:** 10.64898/2026.04.02.26349960

**Authors:** Taiyu Zhu, Alexander Tashevski, Maxime Taquet, Matilda Azis, Tia Jani, Matthew R. Broome, Thomas Kabir, Amedeo Minichino, Graham K. Murray, Matthew M. Nour, Ilina Singh, Paolo Fusar-Poli, Alejo Nevado-Holgado, Philip McGuire, Dominic Oliver

## Abstract

Psychosis prevention relies on early detection of individuals at clinical high risk for psychosis (CHR-P). The effectiveness of the CHR-P state is constrained, in part due to clinical assessments requiring specialist interpretation of narrative interviews, limiting scalability. Here, we evaluate whether large language models (LLMs; deep learning models trained on large text corpora to process and generate language) can extract clinically meaningful information from such interviews to support psychosis risk assessment. We assessed 11 open-weight LLMs on 678 partial PSYCHS interview transcripts from 373 participants (77.7% CHR-P). Models inferred CHR-P status and estimated severity and frequency across 15 symptom domains, benchmarked against researcher-rated scores. Larger models achieved the strongest classification performance (Llama-3.3-70B: accuracy **= 0.80**, sensitivity **= 0.93**, specificity **= 0.58**). LLM-generated symptom scores showed good correlations with researcher-rated scores (ICC**_sev_ = 0.74**, ICC**_freq_ = 0.75**). Performance disparities were minimal across most demographic groups but varied across sites. Generated summaries were largely faithful to source transcripts, with low rates of clinically relevant confabulation (3%). Errors primarily reflected over-pathologisation of non-clinical experiences. While accuracy scaled with model size, smaller models achieved competitive performance with substantially lower computational cost. These findings demonstrate that open-weight LLMs have the potential to assess psychosis risk from psychometric interview transcripts, supporting scalable, human-in-the-loop approaches to early detection.

## Introduction

Psychosis accounts for 10% of the global health burden among young people [1]. In almost 80% of individuals, the first episode of psychosis is preceded by a detectable clinical high risk for psychosis (CHR-P) phase [2, 3], characterized by attenuated psychotic symptoms and functional impairment [4]. Specialized CHR-P services seek to identify people during this window to enable timely monitoring and preventive intervention. Such early detection is clinically meaningful: compared to individuals first identified at the first episode stage, those engaging with CHR-P services have shorter pathways to care and reduced subsequent inpatient use [5]. However, despite this clinical impact, only 5-14% of individuals who later develop psychosis are currently identified during the CHR-P stage, limiting the population-level reach of preventive care [6, 7]. A low detection rate is also observed for first-episode cases identified by early intervention services [8]. This suggests that improving detection and assessment of emerging psychosis (either CHR-P or first episode) is the rate limiting step towards better preventive and early intervention care. Digital health solutions have the potential to help address this gap [9, 10].

While semi-structured CHR-P assessments, such as the Comprehensive Assessment of At-Risk Mental States [CAARMS] [11], Structured Interview for Psychosis-Risk Syndromes [SIPS] [12], Positive SYmptoms and Diagnostic Criteria for the CAARMS Harmonized with the SIPS [PSYCHS] [13], have excellent prognostic accuracy (area-under-the-curve = 0.85) [14], they require highly trained clinicians who have had specialised training in the assessment of subtle symptoms across multiple dimensions, which can take up to two hours per assessment. This represents a major barrier to effective CHR-P detection. Even in well-resourced services, this requires a relatively large amount of clinician time and can lead to delayed or missed detection. In addition, interpretation of symptoms is subjective and can vary across raters and settings, leading to differences in sample ascertainment and monitoring. While detection and CHR-P assessment is recommended by clinical guidelines [15], there is no standard for tools or versions, which exacerbates inconsistencies. This underlines the need for standardized, reproducible and scalable assessments [13, 16].

Large language models (LLMs) are artificial intelligence models that are trained on large text corpora, allowing them to understand and generate human language. Recent advances in LLMs have demonstrated these capabilities across healthcare tasks [17, 18]. LLMs have the potential to standardize documentation of clinically relevant information and expand access to mental healthcare, where assessment depends on nuanced contextual interpretation of language. Recent studies have begun to demonstrate this potential, for example, assessing empathy and adherence to counseling principles in LLM-generated responses [19], and benchmarking aspect-based summarization of psychotherapy sessions to support clinician documentation [20].

Collectively, these advances could enable LLM-augmented CHR-P interviews, where LLMs can automatically score symptom severity and frequency, as well as assess CHR-P criteria, from interview transcripts. This automated approach could alleviate assessment burden in clinical services and reduce inter-site and inter-rater variability in recruitment and symptom scoring for research studies.

To our knowledge, no prior study has evaluated LLMs for structured psychosis risk assessment from psychometric interviews. Here, we systematically evaluated the ability of 11 open weight LLMs applied to psychometric interview transcripts to accurately (i) assess CHR-P criteria; (ii) score severity and frequency of attenuated positive psychotic symptoms; before assessing (iii) algorithmic fairness, (iv) quality of LLM-generated summary reports and systematic failure modes. Finally, we assessed compute-performance trade-offs: a key practical consideration for implementation.

## Results

We evaluated this framework using the Accelerating Medicines Partnership Schizophrenia (AMP-SCZ) dataset [21], comprising 678 partial transcripts from PSYCHS interviews conducted in English. These transcripts corresponded to the first 30 minutes of semi-structured interviews collected across multiple sites and longitudinal timepoints.

A total of 373 participants had available partial transcripts paired with researcher-rated symptom severity and frequency scores. The sample had a mean age of 21.7 years (*s.d.* = 3.8), with 16.2% aged under 18; 63.3% were female, 4.4% non-binary, 18.5% reported a first language other than English and 55.5% were White.

Transcripts were segmented into 15 PSYCHS symptom domains, yielding *n* = 4,691 transcript–domain observations (from *n* = 4,700 complete transcript instances; 9 instances were missing researcher-rated scores, i.e., ground-truth). These were paired with clinician-derived scores and used to evaluate both classification and regression tasks. Unusual Thoughts and Experiences was the most frequently represented symptom domain (12.5%), whereas Somatic Perceptual Abnormalities was the least represented (4.1%). CHR-P participants completed 7.2 (*s.d.* = 5.3) symptom domains, while HC participants completed 13.0 (*s.d.* = 3.8). As these transcripts only contained information related to the first 30 minutes of the interview, participants may only meet CHR-P criteria according to symptom domains not represented within the available text. We therefore re-assessed CHR-P criteria for each participant visit according to the information contained within complete symptom domain transcripts and guidance documentation. 45.8% of participants meet CHR-P criteria on the basis of symptom domains assessed in the transcripts.

As shown in Fig. 1, we evaluated 11 locally-deployed, open-weight LLMs (Llama-3.2-1B-Instruct, gemma-3n-E2B-it, Llama-3.2-3B-Instruct, medgemma-1.5-4b-it, gemma-3n-E4B-it, Llama-3.1-8B-Instruct, Phi-3-medium-4k-instruct, medgemma-27b-it, Qwen3-30B-A3B-Instruct-2507, Llama-3.3-70B-Instruct, Qwen3-Next-80B-A3B-Instruct) using structured reasoning strategies, including chain-of-thought (CoT) prompting [22], to score 15 symptom domains. For each domain, models were instructed to infer severity and frequency scores from the transcript text and then provide a brief, evidence-based rationale summary for the assigned ratings.

**Fig. 1.**
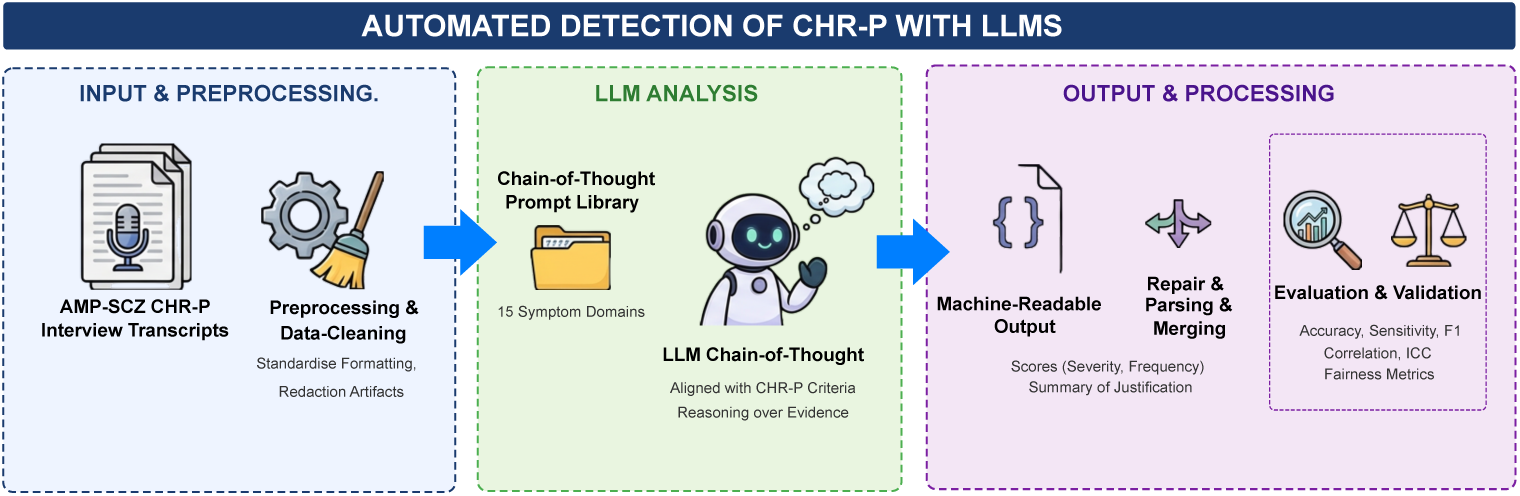
End-to-end pipeline for automated CHR-P symptom scoring from partial interview transcripts. De-identified AMP-SCZ transcripts are preprocessed and analyzed using a library of 15 PSYCHS-aligned prompts. A locally deployed LLM applies CoT to extract symptom evidence and outputs structured JSON containing severity and frequency scores with a brief rationale summary. Outputs are parsed and merged, then subjected to validation and optimization (schema and range checks, consistency checks, and feasibility profiling). Future implementation could support clinical triage and prospective evaluation within CHR-P services.

### CHR-P classification

We first evaluated whether LLM-derived symptom scores could be used to classify CHR-P status at the subject–visit level (*n* = 556). Performance generally improved with model scale (Table 1), with the strongest results achieved by the largest models.

**Table 1.** CHR-P detection performance across open-weight LLMs, grouped by model scale. Metrics are computed at the subject-visit level (*n* = 556) using a pragmatic PSYCHS-aligned gate derived from predicted severity and frequency. Values are proportions.

| Model | Size (B) | Accuracy | Sensitivity | Specificity | Precision | F1 | MCC |
| --- | --- | --- | --- | --- | --- | --- | --- |
| **Small (< 10B)** |  |  |  |  |  |  |  |
| Llama-3.2-1B-Instruct | 1 | 0.599 | 0.819 | 0.227 | 0.641 | 0.719 | 0.056 |
| gemma-3n-E2B-it | 2 | 0.728 | 0.966 | 0.329 | 0.708 | 0.817 | 0.405 |
| Llama-3.2-3B-Instruct | 3 | 0.646 | 1.000 | 0.048 | 0.639 | 0.780 | 0.176 |
| medgemma-1.5-4b-it | 4 | 0.541 | 0.659 | 0.343 | 0.628 | 0.643 | 0.002 |
| gemma-3n-E4B-it | 4 | 0.766 | 0.948 | 0.459 | 0.747 | 0.836 | 0.489 |
| Llama-3.1-8B-Instruct | 8 | 0.745 | 0.960 | 0.382 | 0.724 | 0.825 | 0.442 |
| **Medium (10–30B)** |  |  |  |  |  |  |  |
| Phi-3-medium-4k-instruct | 14 | 0.621 | 0.550 | **0.739** | 0.780 | 0.645 | 0.282 |
| medgemma-27b-it | 27 | 0.750 | 0.911 | 0.478 | 0.746 | 0.821 | 0.445 |
| Qwen3-30B-A3B-Instruct-2507 | 30 | 0.754 | **0.983** | 0.367 | 0.724 | 0.834 | 0.477 |
| **Large (> 30B)** |  |  |  |  |  |  |  |
| Llama-3.3-70B-Instruct | 70 | **0.802** | 0.934 | 0.580 | **0.789** | **0.856** | **0.568** |
| Qwen3-Next-80B-A3B-Instruct | 80 | 0.793 | 0.934 | 0.556 | 0.780 | 0.850 | 0.548 |

Performance improved with model complexity, with the strongest overall results achieved by the largest models. Llama-3.3-70B-Instruct (70 billion parameters) achieved the best overall accuracy (0.802), while maintaining high sensitivity (0.934) and improved specificity (0.580) relative to smaller models. Qwen3-Next-80B-A3B-Instruct (80 billion parameters) performed comparably (accuracy 0.793) with the same sensitivity (0.934) and slightly lower specificity (0.556).

Across models, sensitivity was generally high, consistent with the overestimation tendency observed in Fig. 2 and Supplementary Figs. 1–3, which shifts the operating point toward fewer false negatives at the cost of increased number of false positives. This pattern was most pronounced for smaller models (*<* 10B), with medium-size models (10–30B) generally improving this trade-off.

**Fig. 2.**
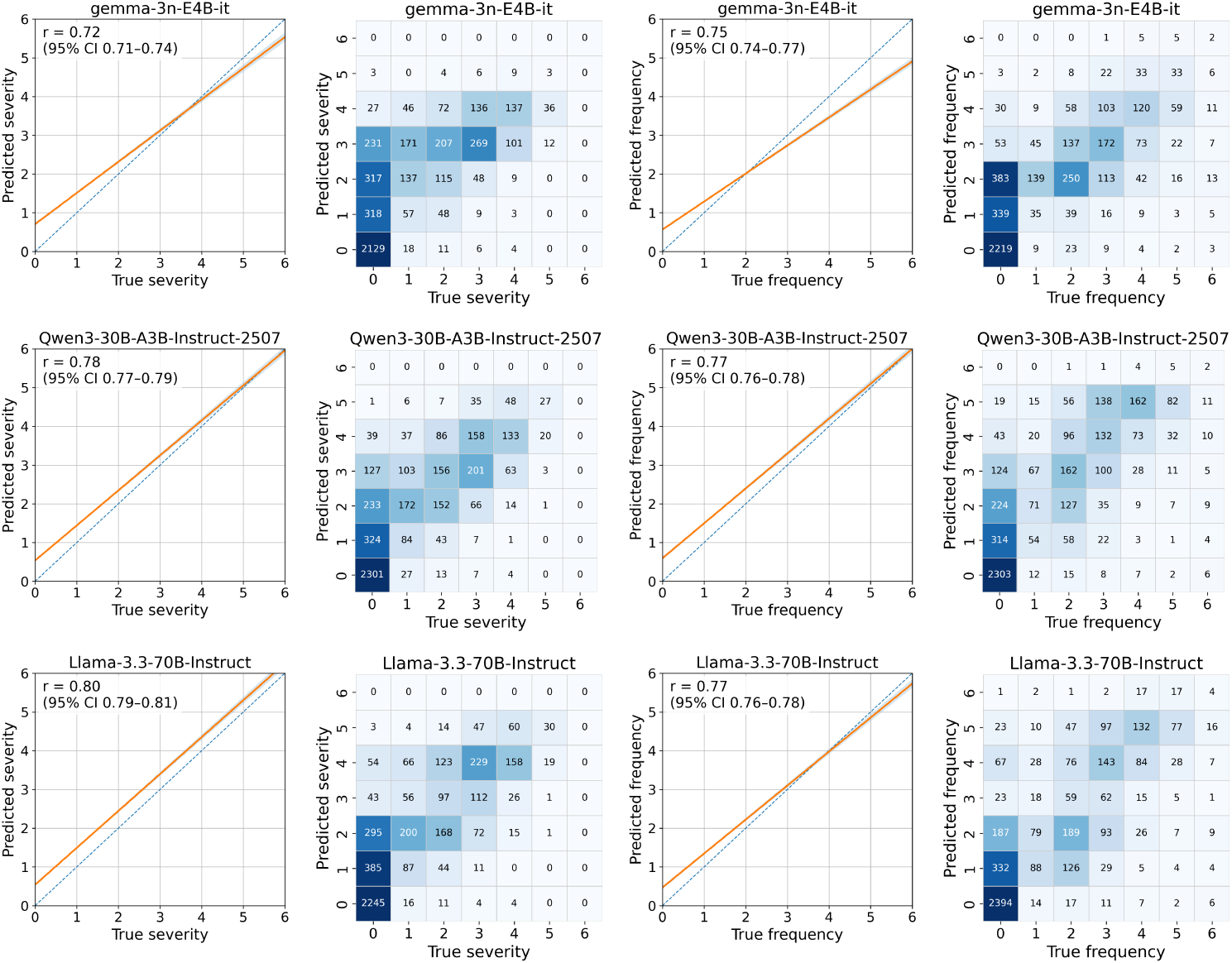
Agreement between LLM-predicted and researcher-rated PSYCHS scores. Three representative models have been selected as the best-performing model within each size tier. Adjacent trend plots show the fitted linear relationship between predicted and researcher-rated scores, with the dashed diagonal representing perfect agreement, the solid line the least-squares regression fit, and the shaded band the 95% prediction interval. Pearson’s correlation coefficient and its 95% confidence interval are shown within each panel. Heatmaps summarise the joint distribution of predicted and researcher-rated (ground-truth) scores for severity/intensity and frequency on the 0–6 scale; each cell is annotated with the number of transcript–item pairs assigned to that score combination. The same colour scale was applied across all heatmaps, with colour intensity capped at 400 counts.

In terms of individual symptom domains, LLMs performed best in scoring severity of Unusual Thoughts and Experiences (Llama-3.3-70B-Instruct: F1= 0.80, MCC= 0.61) and Auditory Perceptual Abnormalities (Llama-3.3-70B-Instruct: F1= 0.77, MCC= 0.76). Performance was difficult to estimate in sparse perceptual-abnormality domains, particularly Gustatory Perceptual Abnormalities, where no positive events were available, and Somatic Perceptual Abnormalities, where only two positive events were available (Supplementary Fig. 4; Supplementary Tables 1–5).

### Scoring symptom severity and frequency

We next evaluated scoring of the PSYCHS symptom domains, comparing LLM-predicted severity and frequency ratings against researcher-rated scores across all available symptom domain transcripts. Table 2 summarizes completeness (the proportion of expected transcript–item pairs yielding valid model outputs), agreement for severity and frequency (Pearson *r*, ICC), and discrimination (AUC). Across models, larger models generally exhibited stronger correlations with researcher-rated scores and more stable output formatting.

**Table 2.** PSYCHS item-level scoring agreement across open-weight LLMs. Metrics were computed at the transcript–item level (*n* = 4,691).

| Model | Size (B) | Completeness |  | Severity |  |  | Frequency |  |  |
| --- | --- | --- | --- | --- | --- | --- | --- | --- | --- |
| | | Raw | Repair | $r$ | ICC | AUC | $r$ | ICC | AUC |
| **Small (< 10B)** |  |  |  |  |  |  |  |  |  |
| Llama-3.2-1B-Instruct | 1 | 0.916 | 1.000 | 0.509 | 0.182 | 0.792 | 0.052 | 0.015 | 0.474 |
| gemma-3n-E2B-it | 2 | 0.998 | 1.000 | 0.694 | 0.627 | 0.884 | 0.711 | 0.702 | 0.890 |
| Llama-3.2-3B-Instruct | 3 | 0.963 | 1.000 | 0.609 | 0.419 | 0.863 | 0.622 | 0.466 | 0.858 |
| medgemma-1.5-4b-it | 4 | 0.730 | 0.808 | 0.408 | 0.198 | 0.771 | 0.610 | 0.592 | 0.888 |
| gemma-3n-E4B-it | 4 | 0.945 | 1.000 | 0.724 | 0.672 | 0.896 | 0.753 | 0.739 | 0.907 |
| Llama-3.1-8B-Instruct | 8 | 0.968 | 0.999 | 0.709 | 0.614 | 0.890 | 0.753 | 0.724 | 0.909 |
| **Medium (10–30B)** |  |  |  |  |  |  |  |  |  |
| Phi-3-medium-4k-instruct | 14 | 0.298 | 0.918 | 0.695 | 0.653 | 0.890 | 0.722 | 0.711 | 0.897 |
| medgemma-27b-it | 27 | 0.926 | 1.000 | 0.736 | 0.689 | 0.891 | 0.756 | 0.749 | 0.903 |
| Qwen3-30B-A3B-Instruct-2507 | 30 | 1.000 | 1.000 | 0.781 | 0.737 | 0.916 | **0.772** | 0.731 | **0.926** |
| **Large (> 30B)** |  |  |  |  |  |  |  |  |  |
| Llama-3.3-70B-Instruct | 70 | **1.000** | 1.000 | **0.797** | 0.743 | **0.920** | 0.770 | 0.748 | 0.911 |
| Qwen3-Next-80B-A3B-Instruct | 80 | 1.000 | 1.000 | 0.784 | **0.767** | 0.912 | 0.770 | **0.749** | 0.913 |

Completeness (Raw) is the proportion of expected transcript-item pairs with valid outputs before recovery; Completeness (Repair) is the proportion after post-processing recovery. Raw output completeness was near ceiling for Qwen3-30B-A3B-Instruct-2507 (99.98%), Llama-3.3-70B-Instruct (100%), and Qwen3-Next-80B-A3B-Instruct (99.98%), indicating that failures were rare under deterministic prompting. By contrast, Phi-3-medium-4k-instruct showed substantially lower raw completeness (29.78%), which increased to 91.81% after automated repair, suggesting that many failures reflected recoverable formatting errors rather than a systematic inability to perform the scoring task. This pattern is consistent with recent structured-output benchmarks [23] showing that Phi-3 models can have comparatively weaker adherence to structured-output formats, including JSON.

Among large models, Llama-3.3-70B-Instruct (ICC_sev_ = 0.743, ICC_freq_ = 0.748) and Qwen3-Next-80B-A3B-Instruct (ICC_sev_ = 0.767; ICC_freq_ = 0.749) performed similarly high. Medium-sized models also performed strongly, with Qwen3-30B-A3B-Instruct-2507 achieving the highest frequency correlation (*r*_freq_ = 0.772). In the small-model group (*<* 10B), gemma-3n-E4B-it provided the strongest overall agreement (ICC_sev_ = 0.672; ICC_freq_ = 0.739).

Predicted scores showed clear concordance with researcher ratings, with most disagreements occurring between adjacent ordinal categories (Fig. 2). However, models exhibited a systematic tendency to overestimate severity and frequency at lower score ranges (Supplementary Figs. 1–3). Meanwhile, both Qwen3-30B-A3B-Instruct-2507 and Llama-3.3-70B-Instruct showed strong correlations with researcher-rated scores for both severity and frequency, with a tendency to overestimate scores when researcher-rated scores were below 4.

In terms of individual symptom domains, LLMs performed best in scoring severity of auditory (Llama-3.3-70B-Instruct: ICC=0.86, *r*=0.88) and visual (Llama-3.3-70B-Instruct: ICC=0.84, *r*=0.87) perceptual abnormalities, with the weakest correlations in scoring severity of erotomanic ideas (Llama-3.3-70B-Instruct: ICC=0.20, *r*=0.34) and somatic perceptual abnormalities (Llama-3.3-70B-Instruct: ICC=0.37, *r*=0.40) (Supplementary Table 1).

### Algorithmic fairness

We evaluated algorithmic fairness at the transcript-domain level using demographic parity (difference in predicted positive rates) and equalised odds (differences in true positive rate (TPR) and false positive rate (FPR)) across age, ethnicity, first language, gender and site. 95%CIs were estimated using 2,000 bootstraps to reflect the variability of fairness metrics (Fig. 3). Results presented in text are from Llama-3.3-70B-Instruct with full results presented in Supplementary Tables 6–11.

**Fig. 3.**
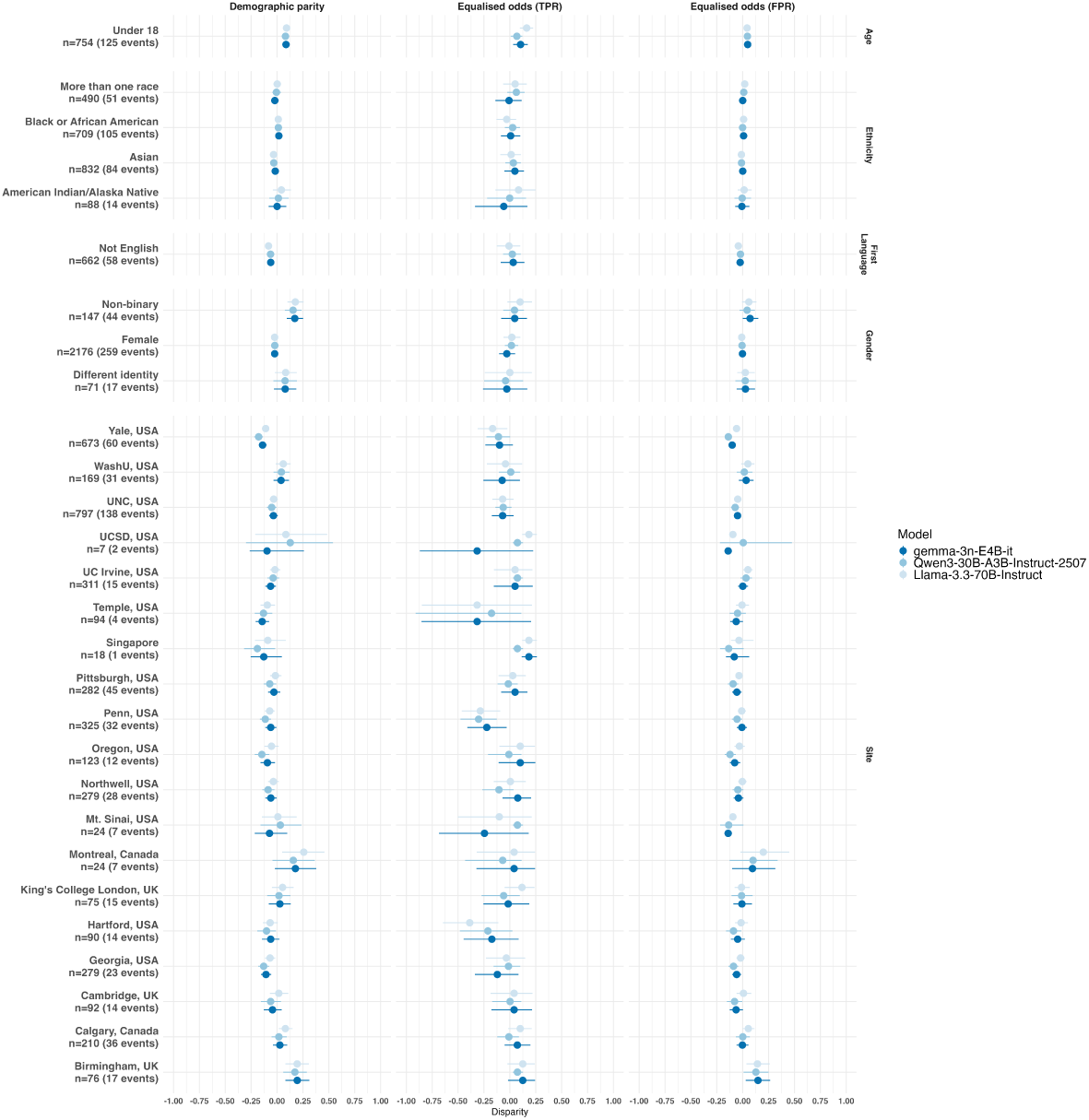
Algorithmic fairness across age (over 18s [reference], under 18s), ethnicity (White [reference], Asian, Black, Mixed, Other), first language (English [reference], Non-English), gender (Male [reference], Female, Non-binary, Different Identity), and site (Melbourne [reference], Birmingham, Calgary, Cambridge, Georgia, Hartford. King’s College London, Montreal, Mt. Sinai, Northwell, Oregon, Penn, Pitt, Singapore, Temple, UC Irvine, UNC, WashU, Yale). Dots represent metric estimates with lines representing bootstrapped 95%CIs. Values of 0 reflect no evident disparities in performance. Wide 95%CIs likely reflect small subgroup sample sizes. Abbreviations: FPR, false positive rate; TPR, true positive rate.

There were small disparities related to age. Comparing under 18s to over 18s, demographic parity difference was 0.010 (95%CIs: 0.06-0.12), TPR disparity was 0.16 (95%CIs: 0.10-0.23) and FPR disparity was 0.04 (95%CIs: 0.02-0.07).

Across ethnicity, disparities were similarly small. Compared to White ethnicity, demographic parity differences ranged from 0.01 (95%CIs: −0.02-0.04) in Black or African American participants to 0.04 (95%CIs: −0.04-0.13) in American Indian/Alaska Native participants, TPR disparities ranged from 0.01 (95%CIs: −0.09-0.11) in Asian participants to 0.08 (95%CIs: −0.15-0.25) in Indian/Alaska Native participants and FPR disparities ranged from 0.01 (95%CIs: −0.02-0.04) in Black or African American participants to 0.02 (95%CIs: −0.01-0.05) in Mixed Race participants.

Similarly, there were small disparities in first language. Comparing participants with English as a first language to those with other languages, demographic parity difference was −0.08 (95%CIs: −0.11 - −0.05). TPR disparity was −0.01 (95%CIs: −0.13-0.11) and FPR disparity was −0.04 (95%CIs: −0.06 - −0.02).

Compared to male participants, disparities were small in female participants (demographic parity difference: −0.02, 95%CIs: −0.05-0.002; TPR disparity: 0.02, 95%CIs: −0.06-0.10; FPR disparity: −0.01, 95%CIs: −0.03-0.01) but more pronounced in non-binary participants (demographic parity difference: 0.18, 95%CIs: 0.10-0.26; TPR disparity: 0.10, 95%CIs: −0.02-0.21; FPR disparity: 0.06, 95%CIs: −0.005-0.13).

Site differences were more heterogeneous. While 84% of sites displayed demographic parity differences ¡0.10 and 89% displayed FPR disparities ¡0.10 compared to the largest site (Melbourne), 58% of sites showed TPR disparities ¿0.10. This was particularly evident in Hartford (−0.38, 95%CIs: −0.66 - −0.11), Temple (−0.32, 95%CIs: −0.85-0.22) and Penn (−0.29, 95%CIs: −0.48 - −0.09).

### Expert-led evaluation of LLM-generated summary reports

Experts (MA and DO) evaluated a randomly selected subset (10%) of summary reports generated by our best performing model (Llama-3.3-70B-Instruct). Expert-generated symptom scores based on the summary reports alone correlated strongly with LLM-generated scores (*r* severity=0.87; *r* frequency=0.87) and researcher-rated scores (*r* severity=0.86; *r* frequency=0.82). LLM-generated and researcher-rated scores were less strongly correlated (*r* severity=0.82; *r* frequency=0.80).

Summary reports were generally accurate, with 93.3% fully representing symptom domain transcript content, while 5.6% had minor inaccuracies and 1.0% with moderate inaccuracies. However, LLM-generated confabulations that impacted scoring were present in 2.7% of reports, generally referring to statements of symptoms being distressing and impacting daily functioning from the scoring guidance, when this had not actually been reported in the transcripts. Moderate omissions (e.g. omitting discussions of substance use related to symptom onset, clarification of experiences being within the range of normal) were present in 0.8% and minor omissions (e.g. clarifications of frequency) in 1.0% of reports. There were no reports that omitted safety concerns raised in the symptom domain transcripts.

Experts (MA and DO) additionally evaluated the 15 cases where model outputs deviated the most from researcher-rated severity and frequency scores to understand systematic patterns of where the model performs poorly. In most of these cases (53%), the LLMs were pathologising experiences that, although unusual, are within the range of typical human experience and/or cultural norms (e.g. feeling mistrustful as a result of a particular adverse experience). In 27% of cases, there was discussion of symptoms outside of the symptom domain being assessed in the interview section, leading to overscoring by the LLMs. In 13% of cases, scores generated by LLMs were considered correct according to the transcript provided, suggesting that researcher-rated scores may use information outside of the transcript. In 7% of cases, LLMs misinterpreted discussion of a situation that led to over-scoring of the symptom domain (e.g. using gesture to communicate in certain instances, or relationship issues).

### Compute-performance trade-offs

We then examined operational trade-offs between model performance and inference cost by relating subject-visit F1 scores to peak GPU memory consumption and token generation speed, as shown in Fig. 4. As expected, performance improved with increasing resource requirements: models achieving the highest F1 scores generally required substantially greater peak GPU memory. Conversely, smaller models clustered at lower memory footprints but delivered more modest F1, consistent with the accuracy limitations observed in the subject-visit evaluation. We additionally quantified the association between F1 score and model size, observing strong positive correlations (Spearman *ρ* = 0.81, *p <* 0.001; Spearman *ρ* = 0.76, *p <* 0.001 after log-transforming model size).

**Fig. 4.**
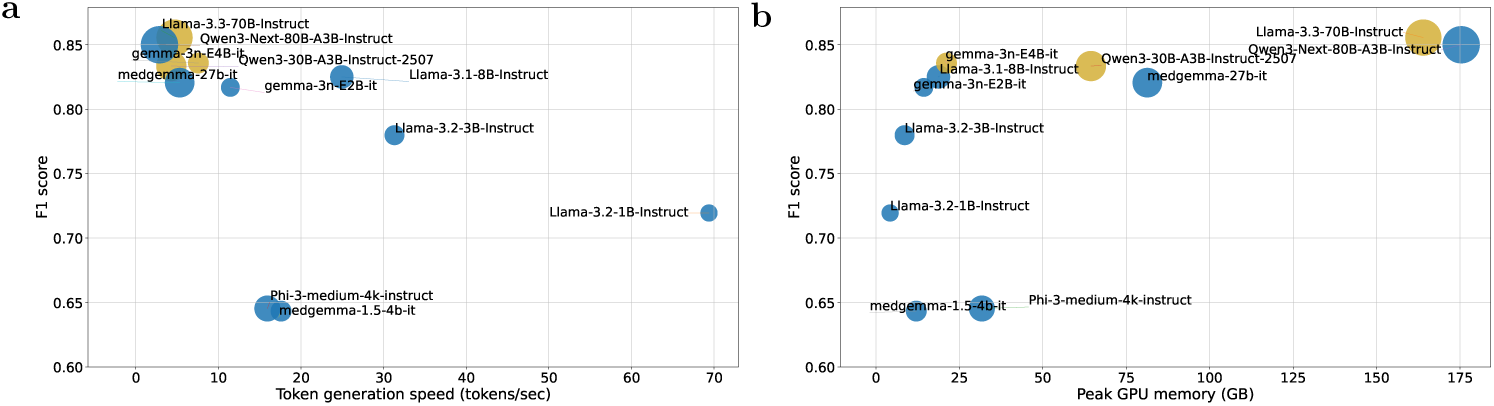
Compute–performance trade-offs across model scales. Scatter plots relate subject–visit CHR-P detection performance (F1 score) to (left) token generation speed (tokens per second) and (right) peak GPU memory consumption and for each evaluated open-weight LLM. Each point represents a model; bubble area is proportional to model parameter count (model scale), and higher values on the y-axis indicate better detection performance.

A similar trade-off was evident for throughput. Models with faster token generation tended to exhibit lower F1, whereas the highest-performing models operated at lower generation speeds (Fig. 4), reflecting an accuracy-latency constraint that is particularly salient for scalable CHR-P triage. Importantly, several mid-sized models occupied an intermediate regime, providing competitive F1 scores at markedly reduced memory demand and improved throughput relative to the largest models. These results motivate selecting model configurations based on the intended clinical operating point, balancing accuracy against hardware availability and acceptable latency for interview-driven workflows.

On the basis of these compute-performance trade-offs, we identified gemma-3n-E4B-it as a particularly practical candidate for deployment. This model achieved a strong subject-visit F1 score while requiring only 21.13 GB peak GPU memory and sustaining approximately 7.59 tokens per second, placing it in a favorable regime of high throughput and low memory demand. These characteristics suggest that gemma-3n-E4B-it could be feasible for a wide range of clinical scenarios with limited GPU capacity, and motivate future work to evaluate efficient deployment pathways, including CPU-oriented serving architectures and, longer term, on-device inference on edge hardware such as smartphones. Importantly, this observation reflects a computational feasibility assessment only and should not be interpreted as a recommendation for immediate clinical deployment of the system.

## Discussion

Our findings demonstrate that open-weight LLMs can extract and score clinically relevant symptomatology from psychosis risk interview transcripts. Across a large, multi-site dataset of interview transcripts, LLMs achieved good classification performance for researcher-rated CHR-P criteria and strong correlations with researcher-rated symptom severity and frequency scores. These findings indicate that contemporary open-weight LLMs can be used to score CHR-P interviews for subtle psychotic symptoms to identify those most likely to reflect people at risk of psychosis, which has potential for increasing scalability of psychosis prevention.

The best-performing models (Llama-3.3-70B-Instruct; Qwen3-Next-80B) achieved good accuracy (80%) for classifying CHR-P status, driven by high sensitivity. This was complemented by accurate symptom severity and frequency scores, with strong correlations with researcher-rated scores (*r ≈* 0.78 *−* 0.80) and similar intraclass correlations to those seen between trained raters (ICC_sev_ = 0.74, ICC_freq_ = 0.75, ICC_CAARMS_*_/_*_SIPS_ = 0.79 *−* 0.88 [11, 24]). These intraclass correlations were lower than those seen for the PSYCHS in the AMP SCZ study (ICC_PSYCHS_ = 0.97), however, this was based on training case vignettes rather than research participants [25]. This performance was attained through structured reasoning strategies with CoT prompting [22]. These findings align with emerging evidence that LLMs can identify the presence of depressive symptoms from interview transcripts [26, 27] and psychiatric diagnoses from interview transcripts [28, 29]. Notably, disagreements were typically confined to adjacent ordinal categories, suggesting that models capture underlying symptom severity even when precise scoring differs. Agreement with researcher-rated scores was strongest in perceptual abnormality domains and lower in less frequently represented or more context-dependent symptom domains. This variability highlights that while LLMs can approximate researcher ratings, performance remains uneven and dependent on both data representation and symptom characteristics. These symptoms are typically challenging to assess without specialist raters [30], who have been trained in the use of psychometric instruments that typically determine service eligibility in clinical settings [31] or inclusion in research studies [21, 32].

Sensitivity was consistently high across models, often at the expense of specificity, reflecting a systematic tendency to overestimate severity and frequency scores. While perfect performance would be ideal, this pattern of performance may be acceptable in a screening context, as delayed identification is associated with longer pathways to care and worse outcomes [5]. In addition, subsequent steps in a stepped assessment framework can allow for more in-depth assessment by a clinician, particularly when given context of known failure modes, to address lower specificity. False positives are not completely neutral and the low specificity of these LLMs could lead to a substantial screening burden. More work is needed to improve the specificity of this framework and any prospective implementation would require calibration according to risk enrichment, local prevalence, service capacity and acceptable burden of false positives.

Algorithmic fairness analyses revealed minimal disparities across age, ethnicity, first language and gender. While still small, there was greater variability in performance across sites. This pattern could reflect heterogeneity in recruitment strategies or interviewing styles, which are known to influence CHR-P ascertainment under human rating conditions [33], as well as the distribution of sample sizes and event rates, which differed substantially across sites. There was no apparent scaling of disparities according to model size, with our best-performing model displaying similar patterns of biases, which suggests that more advanced models may not resolve these disparities alone. Prompting strategies that are tailored to local settings may be an option to address these issues. Our findings reinforce recommendations from the broader LLM ethics literature [17, 18] and mental health roadmaps [34]: the use of LLMs in this context should involve site-level tailoring, prospective auditing and continuous monitoring rather than assuming uniform performance across settings.

Evaluation of summary reports generated by the best-performing model (Llama-3.3-70B-Instruct) suggested that LLM-led summarisation of interview content may provide a clinically meaningful intermediate step to automated scoring. Symptom ratings derived from summaries alone correlated strongly with both LLM-generated and researcher-rated scores, exceeding the direct agreement between LLM and researcher ratings. This suggests that summaries adequately highlight key features from interview content. Overall accuracy was high, with 93.3% of summaries fully representing transcript content and 98.9% showing either full representation or only minor inaccuracies. However, confabulations that affected scoring were present in 3% of reports, typically unsupported claims of distress or functional impairment. As we focused solely on confabulations that affected scoring, this does mean that a larger proportion of summary reports may have contained confabulated inaccuracies that could affect downstream care. Omissions occurred in a minority of cases, particularly around relevant substance use or normalization of experiences. Importantly, no safety concerns were omitted. These findings underscore the promise of LLM-generated summaries for scalable clinical documentation but also highlight the importance of specialist oversight to detect inaccuracies that can have clinically significant impacts on patients’ care.

Several patterns emerged from assessing transcripts where models struggled the most. First, in some scenarios, participants described problems that were within the range of typical experience, which LLMs tended to score as clinically relevant. For example, a participant being concerned about being bullied could be scored high on suspiciousness by an LLM, an understandable response to victimisation in context. Second, pertinent information for one symptom domain may be mentioned in another (e.g. information about hearing voices could be disclosed when responding to questions for unusual thoughts and experiences). In the present study, transcripts were edited according to the sections of the psychometric interview, meaning that this type of information could have been missed. Using LLMs to identify relevant text from the full interview transcript or real-time scoring during the interview could be potential avenues to improve performance in future research.

As expected, model performance increased with model size (*ρ* = 0.81), but at the cost of higher memory consumption and lower token throughput. While larger models typically performed better than smaller ones, it remains to be determined whether their added accuracy justifies the increased requirement for computational resources, mostly in resource-constrained settings. For example, gemma-3n-E4B-it achieved strong performance with modest computational demand, suggesting it may be suitable for services with limited computing resources. Our focus on open-weight models also mitigates privacy concerns associated with transmitting sensitive psychiatric data to external APIs, which is a key ethical consideration in mental health LLM integration [35, 36]. Development of a privacy-preserving route to allow use of frontier models would likely improve performance further than the open-weight models presented here, albeit at the greater computational requirements. The use of frontier models may also enable richer extraction of information from interviews to improve performance. For example, the CHR-P state is characterised not only by positive symptoms, but also by negative symptoms (e.g. blunted affect, alogia, avolition) [37], which may be better detectable through clinical observation during an interview. In our current framework, these features are not well represented because the LLMs used are limited to text-based inputs. More advanced multimodal models could incorporate additional signals from speech (e.g. response latency or vocal characteristics) and video (e.g. facial expressions or body language), potentially allowing for a more comprehensive characterisation of participants’ psychopathology [38].

There are also some limitations to consider. First, the interview transcripts were sourced from a research cohort of CHR-P individuals and healthy controls, which may inflate performance relative to clinical settings where those not meeting CHR-P criteria may exhibit a complex mixture of other psychopathologies. Second, we only had access to partial transcripts from the first 30 minutes of psychometric interviews, meaning that symptom domains in the later stages of the assessment were under-represented, particularly in participants who were more unwell. Sensitivity analyses restricting to complete interviews would therefore be biased towards participants with low symptom endorsement, hindering interpretation of any results. Analysing the full interview transcripts would therefore be a more representative test of the ability of LLMs to assess CHR-P criteria. However, we mitigated this by assessing CHR-P criteria at the symptom domain-level to ensure that results were not skewed by symptoms unassessed in the interview transcripts. Third, fairness analyses were constrained by subgroup sizes and demographic granularity. Moreover, as researcher-rated scores are considered the ground truth in these analyses, apparent fairness may be more reflective of pre-existing systematic biases within these settings (i.e. label bias) rather than true fairness. Fourth, we restricted our analysis to transcripts in the English language so these results may not generalise to interview content in other languages. Fifth, the limited number of available transcripts constrained our ability to perform supervised fine-tuning of the LLMs.

From the LLM perspective, we focused on open-weight instruction-tuned models to maximize adherence to structured output formats. First, we did not evaluate frontier commercial LLMs in this study, given the practical and governance complexities of using externally hosted, remote proprietary models with sensitive mental health data. Second, we also did not assess “reasoning” or “think” LLM variants. Although such models may improve scoring performance, they may also increase the risk of so-called “reasoning hallucinations” [39]. Evaluating these models (for example, GPT-OSS and Qwen “Thinking” variants) therefore represents an important direction for future work in this space. Nevertheless, we also explored retrieval-augmented generation (RAG) by comparing each transcript to 12 exemplar vignettes from the CAARMS manual using cosine similarity, and injecting relevant examples into the prompt when similarity was low to emulate few-shot guidance. This approach did not improve performance and, in some cases, introduced RAG-related confabulations in which models echoed symptoms contained in the retrieved examples rather than the transcript evidence. In this work, we focused on GPT-style, decoder-only LLM architectures rather than BERT-style encoder models. Although encoder-based models can be highly effective for semantic classification, our primary objective was to obtain structured ratings together with a brief, evidence-based summary that supports interpretability and clinician review. Decoder-only LLMs naturally support this joint generation of scores and explanations within a single prompting framework, allowing the scoring decision and its justification to be tightly coupled. This design choice also aligns with the longer-term goal of enabling semi-automated clinical workflows in which models generate provisional ratings and concise summaries that can be efficiently reviewed and adjudicated by clinicians.

Our results demonstrate that open-weight LLMs have the potential to assess psychosis risk from research interview transcripts. While larger LLMs perform best, smaller models may be compute-efficient alternatives. These results highlight the promise of LLM-assisted assessment to improve psychosis prevention at scale, within a human-in-the-loop framework.

## Methods

All analysis followed the Transparent Reporting of a multivariable model for Individual Prognosis or Diagnosis - Large Language Models (TRIPOD-LLM) guidelines [40]) (see reporting checklist).

### Study design and unit of analysis

The primary unit of analysis was a *transcript–domain instance*, defined as a segment of a PSYCHS interview corresponding to one of 15 symptom domains, paired with researcher-rated severity and frequency scores. A total of *n* = 4,691 transcript–domain instances were derived from 556 subject–visit assessments.

### Data source and preprocessing

We used data from the AMP-SCZ initiative (AMP-SCZ Release 3.0), a multi-site international clinical high-risk for psychosis (CHR-P) cohort with harmonized clinical assessments and centralized data processing [21]. Inclusion criteria for participants were: a) Aged 12–30 years inclusive, b) Ability to give informed consent (parental/-guardian consent is obtained for participants aged *<* 18 years), and c) Meeting either CHR-P or healthy control criteria. CHR-P participants met diagnostic criteria for CHR-P determined by the PSYCHS, a harmonized instrument designed to generate attenuated positive psychotic symptom severity and frequency ratings to ascertain diagnostic criteria compatible with both CAARMS and SIPS [11–13]. Healthy controls did not meet CHR-P criteria or have a current or past Cluster A personality disorder, were not receiving any current treatment with psychotropic medication, and did not have a family history (in first-degree relatives) of psychotic spectrum disorders. Exclusion Criteria were a) Antipsychotic medication exposure equivalent to a total lifetime haloperidol dose of *>* 50 mg or current antipsychotic medication at time of baseline assessment, b) Documented history of intellectual disability, c) Past or current clinically relevant central nervous system disorder, d) Traumatic brain injury rated 7 or above on the Traumatic Brain Injury screening instrument,or e) Current or past psychotic disorder. More detail on study design is presented in [21]. Interviews were conducted between June 2022 and January 2025.

All analyses were conducted under existing ethics approvals governing AMP-SCZ data access and use via the NIMH Data Archive (NDA; approved project reference: 23638). In AMP-SCZ, language samples were collected using standardized protocols, including the PSYCHS semi-structured interview, with redacted, partial transcripts of the first 30 minutes of the PSYCHS interview made available through NDA data releases [41, 42]. The PSYCHS is a psychometric interview that assesses 15 distinct positive symptoms categorised into three primary groups: attenuated delusions, attenuated hallucinations, and attenuated thought disorder. The interviews were led by trained research assistants and included a mix of verbatim inquiries and semi-structured follow-up queries. The duration of these interviews varies depending upon participants’ symptoms and communication style, ranging from 30 minutes for those who do not have many symptoms to over two hours for those who experience many psychotic-like symptoms. These interviews were conducted at multiple time points during the study: at screening/baseline and at 1, 2, 3, 6, 12, 18, and 24-month follow-ups. Although the full interview was recorded, only the first 30 minutes were transcribed manually.

Transcription was performed by the HIPAA-compliant transcription service company TranscribeMe! using human transcribers. Transcription was conducted in a “full” verbatim style. This approach captures speech in writing with the greatest possible accuracy, preserving utterances exactly as spoken. As such, the transcripts include filler words, grammatical errors, and nonlinguistic utterances. Each speaker is sequentially labeled (e.g., S1 and S2) according to the order in which they first started to speak in the interview, with S1 typically being a research team member. The transcripts were also detailed with timestamps at the second-level accuracy. The transcript was divided into entries, with each entry representing a change in the speaker. Human editors from TranscribeMe! carefully review the transcripts for protected health information and personally identifiable information. This includes names, geographic details smaller than a state, specific dates related to individuals, contact information, and any unique identifying numbers. An analysis of the AMP SCZ Release 2 data revealed that only 0.17% (SD = 0.0038) of words were redacted, suggesting minimal potential impact on overall results.

We accessed transcripts and corresponding clinical rating variables via the NDA in accordance with AMP-SCZ Data Use Certification and release documentation [43]. CHR-P status, symptom severity scores and symptom frequency scores were measured using PSYCHS. We used de-identified (redacted) partial PSYCHS interview transcripts, paired with researcher-derived severity and frequency scores as well as CHR-P status. From the available AMP-SCZ transcript corpus, we curated a study-specific subset of *n* = 4,691 complete symptom domain transcripts derived from 556 subject–visit assessments, each with matched severity and frequency scores for each item.

Before model inference, transcripts underwent standard preprocessing to improve robustness and comparability across sites and releases. We normalized formatting and speaker-turn structure (interviewer vs participant) and removed common artifacts introduced by transcription and redaction (for example, repeated placeholders and broken delimiters), while preserving the original semantic content and temporal order of the dialogue [41]. Further preprocessing was performed manually by the research team to segment each transcript according to the 15 individual symptom domains, according to the ordering of the PSYCHS interview questions.

Because the available transcripts contained only the first 30 minutes of each interview, some participants may have met CHR-P criteria based on symptom domains not represented in the transcript excerpts. To address this, CHR-P status was re-evaluated for each participant visit using only the symptom domains contained within the available transcripts, in accordance with PSYCHS guidance documentation..

### Symptom scoring task and pipeline overview

Fig 1 summarizes the end-to-end workflow used to produce structured CHR-P symptom scores from transcript text. The scoring task covered 15 symptom domains spanning: (i) unusual thoughts and experiences; (ii) suspiciousness; (iii) unusual somatic ideas; (iv) ideas of guilt; (v) jealous ideas; (vi) unusual religious ideas; (vii) erotomanic ideas; (viii) grandiosity; (ix) auditory; (x) visual; (xi) olfactory; (xii) gustatory (xiii) tactile; and (xiv) somatic perceptual abnormalities; and (xv) disorganized communication/expression. For each domain, the model was required to output an ordinal severity/intensity score and an ordinal frequency score (0–6) consistent with PSYCHS anchors, together with a short evidence-based summary.

Model outputs were returned in a structured format, programmatically parsed and merged across domains, and subjected to automated validation (schema checks, value-range checks, and internal consistency checks) prior to statistical evaluation and feasibility profiling.

### Structured prompting for LLM scoring

We evaluated whether LLMs can infer severity and frequency scores directly from psychometric interview text using inference-time prompting. We constructed a library of 15 domain-specific prompts (see Supplementary Information), one per symptom domain, each specifying: (i) the domain definition and a scoring rubric aligned to the rating scheme used in AMP-SCZ; (ii) anchor descriptions for absent/low/moderate/high endorsement; (iii) instructions to base ratings strictly on explicit evidence in the provided text; and (iv) a strict output schema.

For each transcript–domain instance, the model received the transcript formatted as speaker turns to preserve attribution and conversational context. When symptom-specific segmentation was available, we provided the corresponding segment; otherwise, we provided the full partial transcript and instructed the model to focus only on evidence relevant to the target domain. Prompts included CoT style instructions [22] to encourage systematic evidence identification while constraining the returned output to a machine-readable object.

The required output was a JSON object with three fields: severity, frequency, and summary. Severity and frequency were constrained to integers in the permissible range (0–6). The summary was constrained to 3–5 sentences and required to quote or paraphrase the minimal supporting evidence, explicitly noting uncertainty when evidence was insufficient. An example schema is shown below:

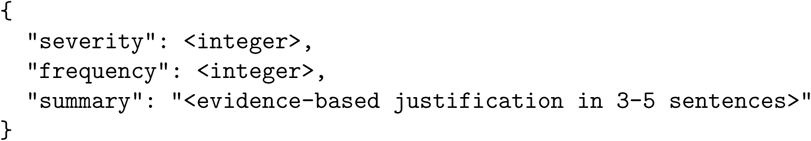

Outputs were programmatically validated for well-formed JSON, presence of required fields, and integer values within the permissible range (0–6). Generations that failed schema validation were retained for post hoc repair. In the analysis pipeline, we attempted to recover severity and frequency values from the returned text using a three-stage fallback procedure. First, we searched for a fenced JSON block (for example, a json{…} block); if parsing succeeded and both scores were valid integers, the output was repaired. If this failed, we searched the full text for the first balanced JSON object, including JSON embedded within other text. If no valid JSON could be extracted, we applied regular-expression patterns to identify numeric values associated with severity and frequency (for example, severity: 4” and frequency: 2”); if both values were recovered and valid, the output was repaired. If all three strategies failed, severity and frequency were left missing for that transcript–item pair.

For each transcript, we applied the relevant prompts across the 15 symptom domains to obtain domain-specific severity and frequency scores and corresponding summaries, and aligned these outputs to researcher-rated scores for evaluation.

### Model deployment and inference configuration

Given the sensitivity of mental health interview data and the requirements of clinical governance, we did not use commercial or closed-source models. Models evaluated included Llama-3.2-1B-Instruct, gemma-3n-E2B-it, Llama-3.2-3B-Instruct, medgemma-1.5-4b-it, gemma-3n-E4B-it, Llama-3.1-8B-Instruct, Phi-3-medium-4k-instruct, medgemma-27b-it, Qwen3-30B-A3B-Instruct-2507, Llama-3.3-70B-Instruct and Qwen3-Next-80B-A3B-Instruct, with parameter sizes ranging from 1B to 80B.

Models were deployed locally on secure institutional infrastructure. Specifically, models were hosted either on King’s College London CREATE HPC with two NVIDIA A100 GPUs (80 GB memory each) or a University of Oxford server equipped with five NVIDIA RTX A6000 GPUs (48 GB memory each). This configuration provided sufficient GPU memory and throughput to support inference with 80B-parameter models, which represented the largest model class evaluated in this study, while keeping all transcript data within the institutional security perimeter. All experiments were conducted in a controlled software environment using Python 3.11.13, PyTorch 2.8.0, and the Hugging Face transformers library (v4.57.1).

All models were executed using the corresponding Hugging Face LLM checkpoints with sampling disabled to maximize reproducibility. We used deterministic decoding (temperature set to 0), such that at each generation step the next token was selected as the most probable token under the model (i.e., greedy decoding). Inference was run with batch size 1, and we issued a single query per transcript–domain instance, ensuring that each output was uniquely determined by the input text and prompt. Models in the small parameter regime were served on a single GPU, whereas larger models were distributed across multiple GPUs to accommodate memory requirements and maintain stable throughput.

### Performance metrics

We quantified agreement between LLM-derived scores and researcher-derived scores using a combination of classification and correlation metrics.

For assessing CHR-P criteria, we first derived binary labels using symptom severity and frequency scores alongside PSYCHS criteria for each symptom domain. If a participant met PSYCHS criteria on any symptom domain with available transcript data, we considered them to meet CHR-P criteria. We then computed accuracy, sensitivity, specificity, precision and F1 from the confusion matrix. We additionally computed Matthew’s Correlation Coefficient. These metrics were computed only when both outcome classes were present in the evaluated subset.

For assessing accuracy of scoring symptom severity and frequency scores, we computed Pearson correlation coefficients (r) and corresponding p-values using pairwise complete observations. Correlations were reported only when at least three paired observations were available. To quantify absolute agreement on an interval scale, we computed the intraclass correlation coefficient ICC(A,1), corresponding to a two-way random-effects model with absolute agreement for single measurements (often reported as ICC2). ICC was estimated by restructuring paired researcher- and model-rated scores into a long format with two raters (researcher vs model) and a unique target index for each paired observation; ICC values were reported only when at least three pairs were available. We additionally computed the area under the receiver operating characteristic curve (AUC).

For algorithmic fairness, we assessed the impact of age (over 18s [reference], under 18s), ethnicity (White [reference], Asian, Black, Mixed, Other), first language (English [reference], other), gender (Male [reference], Female, Non-binary, Different Identity), and site (Melbourne [reference], Birmingham, Calgary, Cambridge, Georgia, Hartford. King’s College London, Montreal, Mt. Sinai, Northwell, Oregon, Penn, Pitt, Singapore, Temple, UC Irvine, UNC, WashU, Yale). For each characteristic, we computed demographic parity and equalised odds (for both true positive rate and false positive rate). 95%CIs were computed through 2,000 bootstraps to provide estimates of variability.

### Evaluating summary reports

We selected 10% of outputs from the best-performing LLM to evaluate, stratifying by symptom domain, severity score and frequency score to maximise generalisability to the full set of transcripts.

Two authors with expertise in CHR-P interviews (MA and DO) reviewed each summary report independently, scoring symptom severity and frequency according to PSYCHS scoring criteria, blinded to LLM-generated and researcher-rated scores. Correlations between scores based on the reports and (i) LLM-generated scores; (ii) researcher-rated scores were calculated using Pearson’s correlation coefficients (r).

In addition, we rated quality of the summary, indexed by accuracy to the transcript (scored 1-5), presence of confabulations impacting scoring, severity of omitted information (scored none, minor, moderate or major) and safety (missed mentions of risks to self or others).

### Hard cases and systematic failure modes

We ranked all symptom domain transcripts according to: Rank A: the most deviated overall (distance across all 11 LLMs) and Rank B: the most deviated relative to the top-performing models across the three model size groups. We selected 15 cases based on the sum of these ranks, representing the largest deviations in scoring.

Two authors with CHR-P interview training (MA and DO) reviewed 15 cases independently and derived failure themes through consensus meetings.

### Compute-performance trade-offs

Compute-performance trade-offs (peak GPU memory and token generation speed) were estimated on NVIDIA RTX A6000 GPUs using a standardized micro-benchmark: 20 subjects were randomly selected and the same set of prompts was executed five times per subject. We recorded peak GPU memory usage and tokens-per-second during generation and summarized these measurements to compare operational feasibility across model scales.

## Data availability

Data presented in this paper are available in the National Institute of Mental Health (NIMH) Data Archive (NDA). On the NDA site ([NIMH Data Archive—AMPSCZ](https://nda.nih.gov/ampscz)), there is information about available data and how it can be obtained.

## Code availability

The analysis code supporting this study is publicly available at https://github.com/tndrg/CHRP_LLM.

## Acknowledgements

MT, TK, AM, IS, PM and DO are supported by the National Institute for Health and Care Research (NIHR) Oxford Health Biomedical Research Centre. GKM is supported by the NIHR Cambridge Biomedical Research Centre (NIHR203312) and the NIHR Applied Research Collaboration East of England. TZ is supported by the National Institute for Health and Care Research (NIHR) Maudsley Biomedical Research Centre (BRC). The views expressed are those of the author(s) and not necessarily those of the NHS, the NIHR or the Department of Health and Social Care. AM is supported by a Wellcome Trust Early Career Award (304693/Z/23/Z). PFP is supported by #NEXTGENERATIONEU (NGEU), funded by the Ministry of University and Research (MUR), National Recovery and Resilience Plan (NRRP), project MNESYS (PE0000006) – A Multiscale integrated approach to the study of the nervous system in health and disease (DN. 1553 11.10.2022).

## Author Contributions

TZ, AN-H, PM and DO conceptualised the study. TZ, AT, AN-H and DO conceptualised the large language model (LLM) pipeline. TZ implemented the LLM pipeline and deployed models. MA and DO performed expert evaluation of summary reports. TZ, TJ and DO accessed and verified the data, and ran the statistical analyses. TZ and DO wrote the initial draft of the manuscript. All authors (TZ, AT, MT, MA, TJ, MRB, TK, AM, GKM, MMN, IS, PF-P, AN-H, PM and DO) drafted, edited and approved the final version of the manuscript.

