## Supplementary Material for "Evaluating Large Language Models for Assessment of Psychosis Risk"

### Supplementary information: Evaluating Assessment of Psychosis Risk Using Large Language Models

#### Score agreement across model scales

Supplementary Figs. 1–3 summarize predicted versus clinician-rated scores for the remaining models, stratified by model scale (small, medium, and large checkpoints). For each model, scatter plots show item-level predicted scores against clinician ratings for severity and frequency, with the identity line indicating perfect agreement and point size reflecting the number of overlapping observations. The accompanying heatmaps display the joint distribution of predicted and true scores, highlighting where models concentrate mass along the diagonal (good agreement) versus systematic off-diagonal shifts (bias).

Across models, agreement was typically strongest near commonly observed rating levels, and most discrepancies reflected off-by-one differences between adjacent categories, consistent with the ordinal nature of PSYCHS scoring. Smaller models more frequently exhibited upward shifts at lower clinician ratings, consistent with a tendency toward overestimation and reduced specificity in downstream detection, whereas larger models showed tighter clustering around the identity line and more concentrated diagonal structure in the heatmaps. Together, these figures provide a scale-stratified view of calibration and error patterns that complements the aggregated metrics reported in Table 2.

#### CHR-P detection performance stratified by symptom domain

Supplementary Fig. 4 summarizes domain-stratified CHR-P detection accuracy for three representative models (gemma-3n-E4B-it, Qwen3-30B-A3B-Instruct-2507, and Llama-3.3-70B-Instruct), illustrating how performance varies across symptom domains. Supplementary Tables 1–5 show domain-stratified CHR-P detection performance across the Llama, Qwen, Gemma, MedGemma, and Phi model families. For each symptom domain, we report the number of subject-visits and positive events, alongside accuracy, sensitivity, specificity, F1 score, and MCC. These results were calculated at the subject-visit level using a pragmatic PSYCHS-aligned detection rule derived from the predicted severity and frequency scores, allowing comparison of model performance across different symptom domains and model families.

#### Fairness diagnostics: demographic parity and equalized odds

Supplementary Table 6 reports group-stratified performance to assess fairness-related properties of CHR-P detection across sex, race, and recruitment site. We summarize the predicted positive rate (PPR), the true positive rate (TPR; sensitivity), and the false positive rate (FPR; 1–specificity) for each subgroup. PPR provides a descriptive check of demographic parity by quantifying how frequently the model assigns a positive CHR-P classification within each group, whereas TPR and FPR jointly inform equalized odds by indicating whether error rates are comparable across groups.

These stratified summaries are intended as diagnostic measures rather than definitive fairness claims. Differences in PPR, TPR, or FPR can reflect a mixture of factors, including heterogeneity in case mix, transcript completeness, and site-specific interviewing practices, as well as potential model bias. Accordingly, we interpret subgroup deviations as signals motivating further prospective evaluation, including larger samples per subgroup, confidence intervals for group metrics, and clinician-led review of discordant cases to understand sources of disparity before any clinical deployment.

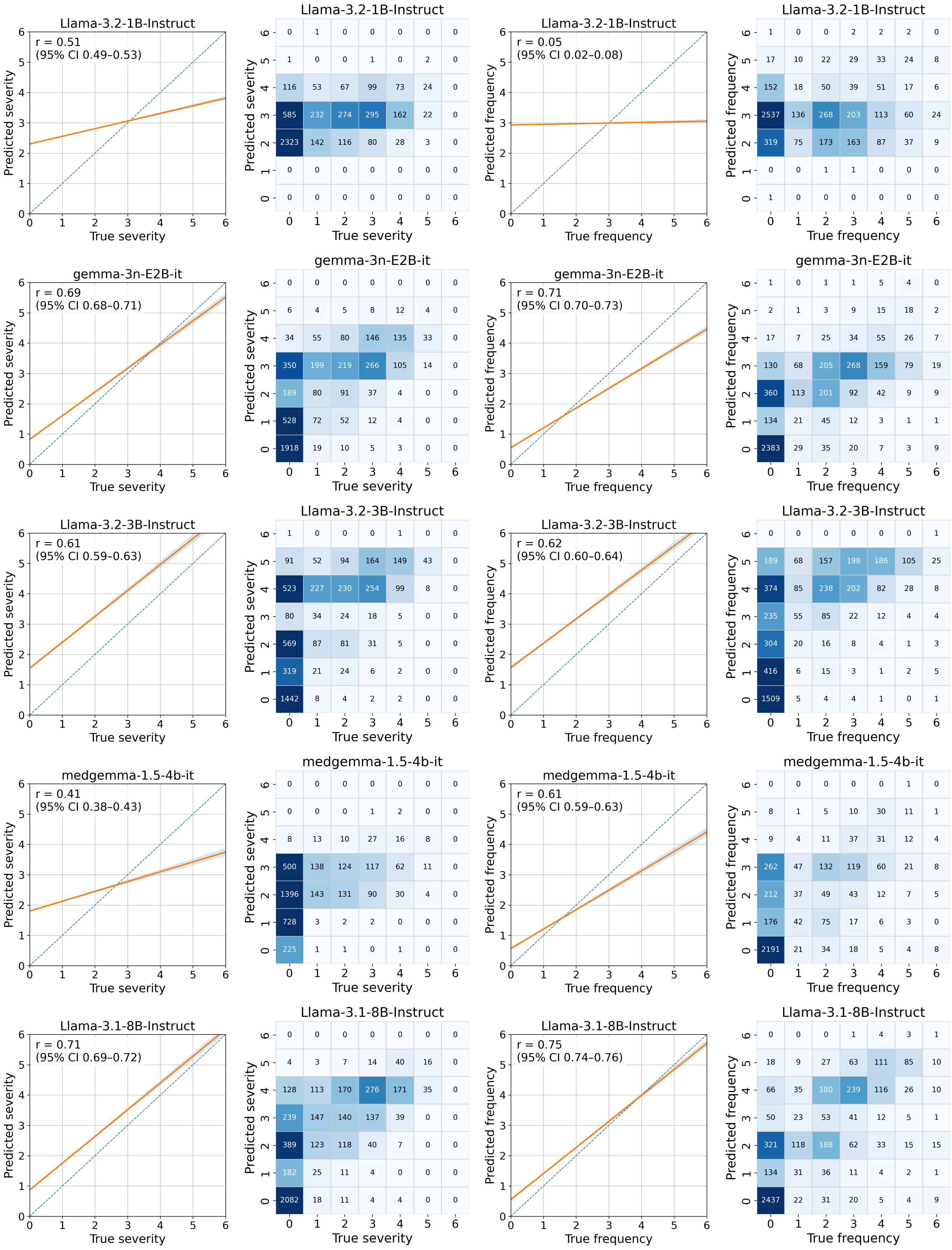


**Supplementary Figure 1:** Score agreement for small models (< 10B).

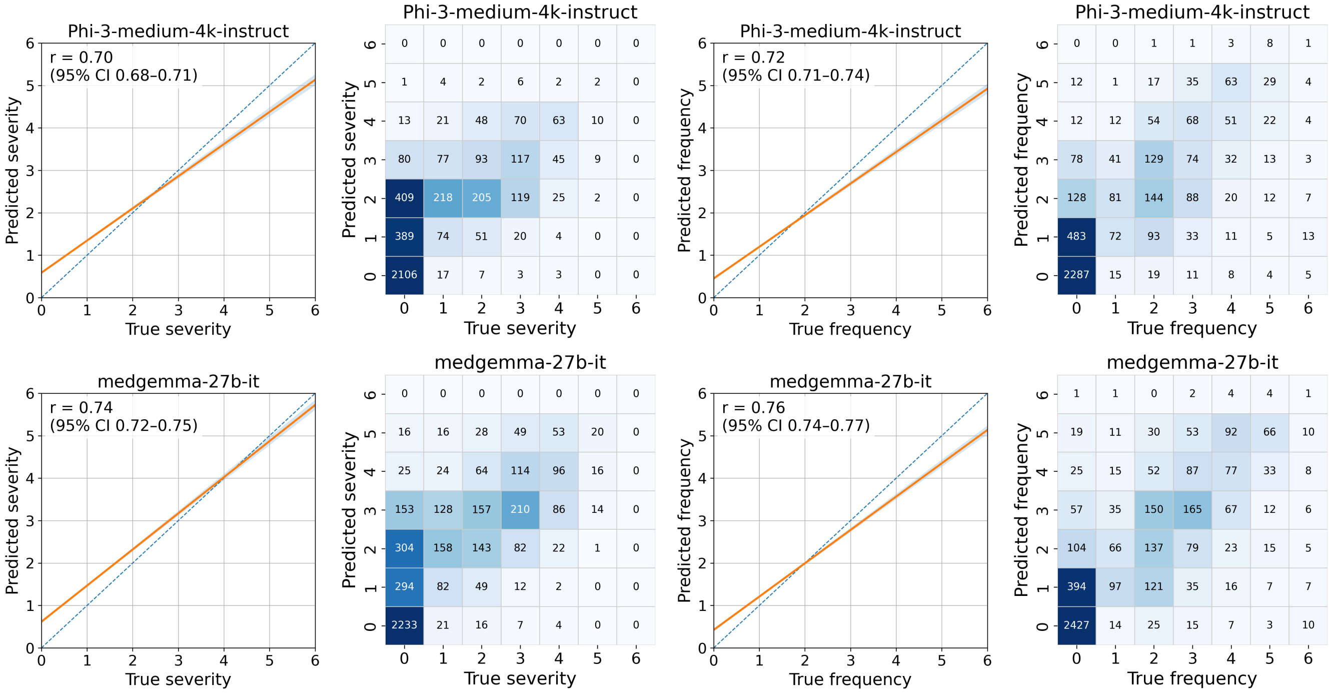


**Supplementary Figure 2:** Score agreement for medium models (10–30B).

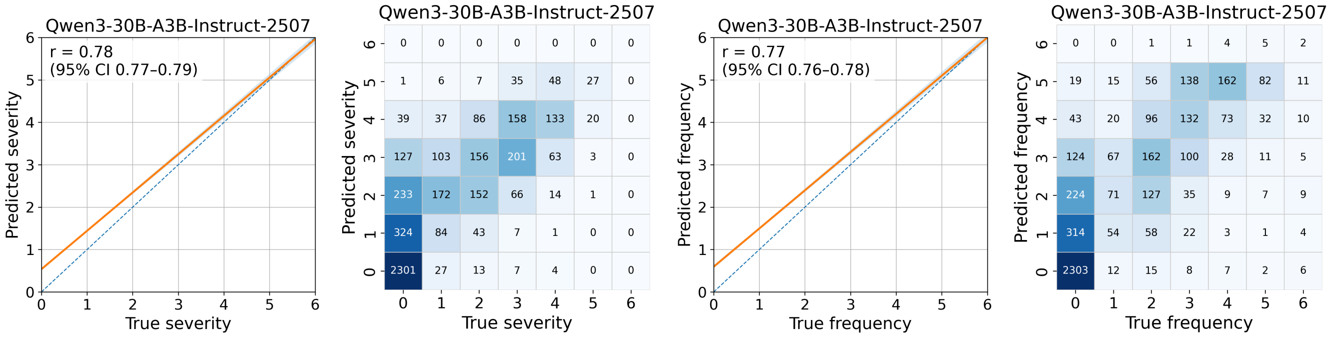


**Supplementary Figure 3:** Score agreement for large models ($> 30$B).

**Supplementary Table 1:** CHR-P detection performance across the Llama family, stratified by symptom domain. Metrics are computed at the subject-visit level ($n = 556$) using a pragmatic PSYCHS-aligned gate derived from predicted severity and frequency. Values are proportions.

| Model | Symptom Domain | n (Events) | Accuracy | Sensitivity | Specificity | F1 | MCC |
|---|---|---|---|---|---|---|---|
| Llama-3.2-1B-Instruct | Q1 | 552 (255) | 0.612 | 0.475 | 0.731 | 0.531 | 0.213 |
| Llama-3.2-1B-Instruct | Q2 | 464 (165) | 0.741 | 0.503 | 0.873 | 0.580 | 0.410 |
| Llama-3.2-1B-Instruct | Q3 | 427 (38) | 0.707 | 0.868 | 0.692 | 0.346 | 0.332 |
| Llama-3.2-1B-Instruct | Q4 | 386 (55) | 0.728 | 0.709 | 0.731 | 0.426 | 0.327 |
| Llama-3.2-1B-Instruct | Q5 | 352 (5) | 0.722 | 0.600 | 0.723 | 0.058 | 0.085 |
| Llama-3.2-1B-Instruct | Q6 | 334 (7) | 0.943 | 0.286 | 0.957 | 0.174 | 0.163 |
| Llama-3.2-1B-Instruct | Q7 | 327 (3) | 0.713 | 0.333 | 0.716 | 0.021 | 0.010 |
| Llama-3.2-1B-Instruct | Q8 | 302 (6) | 0.616 | 0.833 | 0.611 | 0.079 | 0.127 |
| Llama-3.2-1B-Instruct | Q9 | 261 (26) | 0.854 | 0.692 | 0.872 | 0.486 | 0.437 |
| Llama-3.2-1B-Instruct | Q10 | 237 (26) | 0.886 | 0.577 | 0.924 | 0.526 | 0.464 |
| Llama-3.2-1B-Instruct | Q11 | 221 (6) | 0.919 | 0.667 | 0.926 | 0.308 | 0.335 |
| Llama-3.2-1B-Instruct | Q12 | 218 (0) | NA | NA | NA | NA | NA |
| Llama-3.2-1B-Instruct | Q13 | 215 (6) | 0.921 | 0.500 | 0.933 | 0.261 | 0.264 |
| Llama-3.2-1B-Instruct | Q14 | 209 (2) | 0.971 | 0.000 | 0.981 | NA | -0.014 |
| Llama-3.2-1B-Instruct | Q15 | 195 (10) | 0.569 | 0.700 | 0.562 | 0.143 | 0.116 |
| Llama-3.2-1B-Instruct | Overall | 4700 (610) | 0.766 | 0.548 | 0.798 | 0.378 | 0.270 |
| Llama-3.2-1B-Instruct | Subject-level Overall | 556 (349) | 0.599 | 0.819 | 0.227 | 0.719 | 0.056 |
| Llama-3.2-3B-Instruct | Q1 | 552 (255) | 0.714 | 0.961 | 0.502 | 0.756 | 0.509 |
| Llama-3.2-3B-Instruct | Q2 | 464 (165) | 0.619 | 0.982 | 0.418 | 0.647 | 0.428 |
| Llama-3.2-3B-Instruct | Q3 | 427 (38) | 0.609 | 0.921 | 0.578 | 0.295 | 0.285 |
| Llama-3.2-3B-Instruct | Q4 | 386 (55) | 0.528 | 0.964 | 0.456 | 0.368 | 0.300 |
| Llama-3.2-3B-Instruct | Q5 | 352 (5) | 0.676 | 1.000 | 0.671 | 0.081 | 0.168 |
| Llama-3.2-3B-Instruct | Q6 | 334 (7) | 0.853 | 0.857 | 0.853 | 0.197 | 0.276 |
| Llama-3.2-3B-Instruct | Q7 | 327 (3) | 0.278 | 0.667 | 0.275 | 0.017 | -0.013 |
| Llama-3.2-3B-Instruct | Q8 | 302 (6) | 0.662 | 0.667 | 0.662 | 0.073 | 0.097 |
| Llama-3.2-3B-Instruct | Q9 | 261 (26) | 0.782 | 1.000 | 0.757 | 0.477 | 0.487 |
| Llama-3.2-3B-Instruct | Q10 | 237 (26) | 0.743 | 0.885 | 0.725 | 0.430 | 0.402 |
| Llama-3.2-3B-Instruct | Q11 | 221 (6) | 0.919 | 1.000 | 0.916 | 0.400 | 0.479 |
| Llama-3.2-3B-Instruct | Q12 | 218 (0) | NA | NA | NA | NA | NA |
| Llama-3.2-3B-Instruct | Q13 | 215 (6) | 0.744 | 1.000 | 0.737 | 0.179 | 0.269 |
| Llama-3.2-3B-Instruct | Q14 | 209 (2) | 0.895 | 1.000 | 0.894 | 0.154 | 0.273 |
| Llama-3.2-3B-Instruct | Q15 | 195 (10) | 0.621 | 0.800 | 0.611 | 0.178 | 0.184 |
| Llama-3.2-3B-Instruct | Overall | 4700 (610) | 0.684 | 0.956 | 0.644 | 0.440 | 0.406 |
| Llama-3.2-3B-Instruct | Subject-level Overall | 556 (349) | 0.646 | 1.000 | 0.048 | 0.780 | 0.176 |
| Llama-3.1-8B-Instruct | Q1 | 552 (255) | 0.803 | 0.863 | 0.751 | 0.801 | 0.613 |
| Llama-3.1-8B-Instruct | Q2 | 464 (165) | 0.759 | 0.897 | 0.682 | 0.725 | 0.555 |
| Llama-3.1-8B-Instruct | Q3 | 427 (38) | 0.761 | 0.895 | 0.748 | 0.400 | 0.396 |
| Llama-3.1-8B-Instruct | Q4 | 386 (55) | 0.731 | 0.891 | 0.704 | 0.485 | 0.428 |
| Llama-3.1-8B-Instruct | Q5 | 352 (5) | 0.855 | 1.000 | 0.853 | 0.164 | 0.276 |
| Llama-3.1-8B-Instruct | Q6 | 334 (7) | 0.979 | 0.714 | 0.985 | 0.588 | 0.588 |
| Llama-3.1-8B-Instruct | Q7 | 327 (3) | 0.884 | 0.333 | 0.889 | 0.050 | 0.067 |
| Llama-3.1-8B-Instruct | Q8 | 302 (6) | 0.940 | 0.167 | 0.956 | 0.100 | 0.081 |
| Llama-3.1-8B-Instruct | Q9 | 261 (26) | 0.877 | 0.923 | 0.872 | 0.600 | 0.588 |
| Llama-3.1-8B-Instruct | Q10 | 237 (26) | 0.878 | 0.769 | 0.891 | 0.580 | 0.535 |
| Llama-3.1-8B-Instruct | Q11 | 221 (6) | 0.955 | 1.000 | 0.953 | 0.545 | 0.598 |
| Llama-3.1-8B-Instruct | Q12 | 218 (0) | NA | NA | NA | NA | NA |
| Llama-3.1-8B-Instruct | Q13 | 215 (6) | 0.926 | 1.000 | 0.923 | 0.429 | 0.502 |
| Llama-3.1-8B-Instruct | Q14 | 209 (2) | 0.967 | 0.000 | 0.976 | NA | -0.015 |
| Llama-3.1-8B-Instruct | Q15 | 195 (10) | 0.800 | 0.500 | 0.816 | 0.204 | 0.174 |
| Llama-3.1-8B-Instruct | Overall | 4700 (610) | 0.855 | 0.859 | 0.855 | 0.606 | 0.563 |
| Llama-3.1-8B-Instruct | Subject-level Overall | 556 (349) | 0.745 | 0.960 | 0.382 | 0.825 | 0.442 |
| Llama-3.3-70B-Instruct | Q1 | 552 (255) | 0.806 | 0.816 | 0.798 | 0.795 | 0.612 |
| Llama-3.3-70B-Instruct | Q2 | 464 (165) | 0.795 | 0.806 | 0.789 | 0.737 | 0.577 |
| Llama-3.3-70B-Instruct | Q3 | 427 (38) | 0.834 | 0.684 | 0.848 | 0.423 | 0.380 |
| Llama-3.3-70B-Instruct | Q4 | 386 (55) | 0.788 | 0.800 | 0.785 | 0.518 | 0.447 |
| Llama-3.3-70B-Instruct | Q5 | 352 (5) | 0.943 | 0.200 | 0.954 | 0.091 | 0.085 |
| Llama-3.3-70B-Instruct | Q6 | 334 (7) | 0.979 | 0.429 | 0.991 | 0.462 | 0.452 |
| Llama-3.3-70B-Instruct | Q7 | 327 (3) | 0.972 | 0.333 | 0.978 | 0.182 | 0.192 |
| Llama-3.3-70B-Instruct | Q8 | 302 (6) | 0.970 | 0.167 | 0.986 | 0.182 | 0.167 |
| Llama-3.3-70B-Instruct | Q9 | 261 (26) | 0.946 | 0.923 | 0.949 | 0.774 | 0.757 |
| Llama-3.3-70B-Instruct | Q10 | 237 (26) | 0.928 | 0.654 | 0.962 | 0.667 | 0.627 |
| Llama-3.3-70B-Instruct | Q11 | 221 (6) | 0.977 | 0.833 | 0.981 | 0.667 | 0.670 |
| Llama-3.3-70B-Instruct | Q12 | 218 (0) | NA | NA | NA | NA | NA |
| Llama-3.3-70B-Instruct | Q13 | 215 (6) | 0.967 | 0.667 | 0.976 | 0.533 | 0.529 |
| Llama-3.3-70B-Instruct | Q14 | 209 (2) | 0.981 | 0.000 | 0.990 | NA | -0.010 |
| Llama-3.3-70B-Instruct | Q15 | 195 (10) | 0.841 | 0.500 | 0.859 | 0.244 | 0.217 |
| Llama-3.3-70B-Instruct | Overall | 4700 (610) | 0.898 | 0.774 | 0.916 | 0.663 | 0.613 |
| Llama-3.3-70B-Instruct | Subject-level Overall | 556 (349) | 0.802 | 0.934 | 0.580 | 0.856 | 0.568 |

**Supplementary Table 2:** CHR-P detection performance across the Qwen family, stratified by symptom domain. Metrics are computed at the subject-visit level ($n = 556$) using a pragmatic PSYCHS-aligned gate derived from predicted severity and frequency. Values are proportions.

| Model | Symptom Domain | n (Events) | Accuracy | Sensitivity | Specificity | F1 | MCC |
|---|---|---|---|---|---|---|---|
| Qwen3-30B-A3B-Instruct-2507 | Q1 | 552 (255) | 0.775 | 0.929 | 0.643 | 0.793 | 0.588 |
| Qwen3-30B-A3B-Instruct-2507 | Q2 | 464 (165) | 0.750 | 0.885 | 0.676 | 0.716 | 0.537 |
| Qwen3-30B-A3B-Instruct-2507 | Q3 | 427 (38) | 0.799 | 0.763 | 0.802 | 0.403 | 0.373 |
| Qwen3-30B-A3B-Instruct-2507 | Q4 | 386 (55) | 0.764 | 0.836 | 0.752 | 0.503 | 0.437 |
| Qwen3-30B-A3B-Instruct-2507 | Q5 | 352 (5) | 0.881 | 0.800 | 0.882 | 0.160 | 0.242 |
| Qwen3-30B-A3B-Instruct-2507 | Q6 | 334 (7) | 0.964 | 0.857 | 0.966 | 0.500 | 0.537 |
| Qwen3-30B-A3B-Instruct-2507 | Q7 | 327 (3) | 0.905 | 0.333 | 0.910 | 0.061 | 0.081 |
| Qwen3-30B-A3B-Instruct-2507 | Q8 | 302 (6) | 0.924 | 0.167 | 0.939 | 0.080 | 0.061 |
| Qwen3-30B-A3B-Instruct-2507 | Q9 | 261 (26) | 0.893 | 0.962 | 0.885 | 0.641 | 0.635 |
| Qwen3-30B-A3B-Instruct-2507 | Q10 | 237 (26) | 0.903 | 0.846 | 0.910 | 0.657 | 0.625 |
| Qwen3-30B-A3B-Instruct-2507 | Q11 | 221 (6) | 0.964 | 1.000 | 0.963 | 0.600 | 0.642 |
| Qwen3-30B-A3B-Instruct-2507 | Q12 | 218 (0) | NA | NA | NA | NA | NA |
| Qwen3-30B-A3B-Instruct-2507 | Q13 | 215 (6) | 0.944 | 0.833 | 0.947 | 0.455 | 0.490 |
| Qwen3-30B-A3B-Instruct-2507 | Q14 | 209 (2) | 0.981 | 0.500 | 0.986 | 0.333 | 0.345 |
| Qwen3-30B-A3B-Instruct-2507 | Q15 | 195 (10) | 0.764 | 0.500 | 0.778 | 0.179 | 0.145 |
| Qwen3-30B-A3B-Instruct-2507 | Overall | 4700 (610) | 0.861 | 0.875 | 0.859 | 0.621 | 0.581 |
| Qwen3-30B-A3B-Instruct-2507 | Subject-level Overall | 556 (349) | 0.754 | 0.983 | 0.367 | 0.834 | 0.477 |
| Qwen3-Next-80B-A3B-Instruct | Q1 | 552 (255) | 0.804 | 0.827 | 0.785 | 0.796 | 0.610 |
| Qwen3-Next-80B-A3B-Instruct | Q2 | 464 (165) | 0.800 | 0.788 | 0.806 | 0.737 | 0.579 |
| Qwen3-Next-80B-A3B-Instruct | Q3 | 427 (38) | 0.843 | 0.632 | 0.864 | 0.417 | 0.367 |
| Qwen3-Next-80B-A3B-Instruct | Q4 | 386 (55) | 0.785 | 0.836 | 0.776 | 0.526 | 0.463 |
| Qwen3-Next-80B-A3B-Instruct | Q5 | 352 (5) | 0.923 | 0.600 | 0.928 | 0.182 | 0.231 |
| Qwen3-Next-80B-A3B-Instruct | Q6 | 334 (7) | 0.982 | 0.857 | 0.985 | 0.667 | 0.676 |
| Qwen3-Next-80B-A3B-Instruct | Q7 | 327 (3) | 0.948 | 0.333 | 0.954 | 0.105 | 0.127 |
| Qwen3-Next-80B-A3B-Instruct | Q8 | 302 (6) | 0.964 | 0.167 | 0.980 | 0.154 | 0.136 |
| Qwen3-Next-80B-A3B-Instruct | Q9 | 261 (26) | 0.923 | 0.923 | 0.923 | 0.706 | 0.690 |
| Qwen3-Next-80B-A3B-Instruct | Q10 | 237 (26) | 0.928 | 0.577 | 0.972 | 0.638 | 0.603 |
| Qwen3-Next-80B-A3B-Instruct | Q11 | 221 (6) | 0.959 | 0.833 | 0.963 | 0.526 | 0.550 |
| Qwen3-Next-80B-A3B-Instruct | Q12 | 218 (0) | NA | NA | NA | NA | NA |
| Qwen3-Next-80B-A3B-Instruct | Q13 | 215 (6) | 0.953 | 0.833 | 0.957 | 0.500 | 0.528 |
| Qwen3-Next-80B-A3B-Instruct | Q14 | 209 (2) | 0.981 | 0.500 | 0.986 | 0.333 | 0.345 |
| Qwen3-Next-80B-A3B-Instruct | Q15 | 195 (10) | 0.841 | 0.400 | 0.865 | 0.205 | 0.164 |
| Qwen3-Next-80B-A3B-Instruct | Overall | 4700 (610) | 0.892 | 0.780 | 0.909 | 0.652 | 0.601 |
| Qwen3-Next-80B-A3B-Instruct | Subject-level Overall | 556 (349) | 0.793 | 0.934 | 0.556 | 0.850 | 0.548 |

**Supplementary Table 3:** CHR-P detection performance across the Gemma family, stratified by symptom domain. Metrics are computed at the subject-visit level ($n = 556$) using a pragmatic PSYCHS-aligned gate derived from predicted severity and frequency. Values are proportions.

| Model | Symptom Domain | n (Events) | Accuracy | Sensitivity | Specificity | F1 | MCC |
|---|---|---|---|---|---|---|---|
| gemma-3n-E2B-it | Q1 | 552 (255) | 0.757 | 0.871 | 0.660 | 0.768 | 0.537 |
| gemma-3n-E2B-it | Q2 | 464 (165) | 0.782 | 0.818 | 0.763 | 0.728 | 0.560 |
| gemma-3n-E2B-it | Q3 | 427 (38) | 0.778 | 0.816 | 0.774 | 0.395 | 0.374 |
| gemma-3n-E2B-it | Q4 | 386 (55) | 0.723 | 0.855 | 0.701 | 0.468 | 0.400 |
| gemma-3n-E2B-it | Q5 | 352 (5) | 0.875 | 0.400 | 0.882 | 0.083 | 0.102 |
| gemma-3n-E2B-it | Q6 | 334 (7) | 0.928 | 0.714 | 0.933 | 0.294 | 0.340 |
| gemma-3n-E2B-it | Q7 | 327 (3) | 0.890 | 0.333 | 0.895 | 0.053 | 0.070 |
| gemma-3n-E2B-it | Q8 | 302 (6) | 0.917 | 0.500 | 0.926 | 0.194 | 0.216 |
| gemma-3n-E2B-it | Q9 | 261 (26) | 0.920 | 0.885 | 0.923 | 0.687 | 0.665 |
| gemma-3n-E2B-it | Q10 | 237 (26) | 0.890 | 0.577 | 0.929 | 0.536 | 0.475 |
| gemma-3n-E2B-it | Q11 | 221 (6) | 0.941 | 1.000 | 0.940 | 0.480 | 0.545 |
| gemma-3n-E2B-it | Q12 | 218 (0) | NA | NA | NA | NA | NA |
| gemma-3n-E2B-it | Q13 | 215 (6) | 0.907 | 0.667 | 0.914 | 0.286 | 0.315 |
| gemma-3n-E2B-it | Q14 | 209 (2) | 0.967 | 0.000 | 0.976 | NA | -0.015 |
| gemma-3n-E2B-it | Q15 | 195 (10) | 0.749 | 0.700 | 0.751 | 0.222 | 0.224 |
| gemma-3n-E2B-it | Overall | 4700 (610) | 0.849 | 0.821 | 0.854 | 0.586 | 0.536 |
| gemma-3n-E2B-it | Subject-level Overall | 556 (349) | 0.728 | 0.966 | 0.329 | 0.817 | 0.405 |
| gemma-3n-E4B-it | Q1 | 552 (255) | 0.774 | 0.827 | 0.727 | 0.771 | 0.554 |
| gemma-3n-E4B-it | Q2 | 464 (165) | 0.782 | 0.788 | 0.779 | 0.720 | 0.550 |
| gemma-3n-E4B-it | Q3 | 427 (38) | 0.808 | 0.763 | 0.812 | 0.414 | 0.384 |
| gemma-3n-E4B-it | Q4 | 386 (55) | 0.780 | 0.800 | 0.776 | 0.509 | 0.437 |
| gemma-3n-E4B-it | Q5 | 352 (5) | 0.938 | 0.400 | 0.945 | 0.154 | 0.172 |
| gemma-3n-E4B-it | Q6 | 334 (7) | 0.964 | 0.714 | 0.969 | 0.455 | 0.473 |
| gemma-3n-E4B-it | Q7 | 327 (3) | 0.960 | 0.333 | 0.966 | 0.133 | 0.152 |
| gemma-3n-E4B-it | Q8 | 302 (6) | 0.957 | 0.167 | 0.973 | 0.133 | 0.115 |
| gemma-3n-E4B-it | Q9 | 261 (26) | 0.920 | 0.885 | 0.923 | 0.687 | 0.665 |
| gemma-3n-E4B-it | Q10 | 237 (26) | 0.924 | 0.577 | 0.967 | 0.625 | 0.586 |
| gemma-3n-E4B-it | Q11 | 221 (6) | 0.959 | 1.000 | 0.958 | 0.571 | 0.619 |
| gemma-3n-E4B-it | Q12 | 218 (0) | NA | NA | NA | NA | NA |
| gemma-3n-E4B-it | Q13 | 215 (6) | 0.935 | 0.500 | 0.947 | 0.300 | 0.299 |
| gemma-3n-E4B-it | Q14 | 209 (2) | 0.981 | 0.000 | 0.990 | NA | -0.010 |
| gemma-3n-E4B-it | Q15 | 195 (10) | 0.795 | 0.600 | 0.805 | 0.231 | 0.218 |
| gemma-3n-E4B-it | Overall | 4700 (610) | 0.880 | 0.780 | 0.895 | 0.628 | 0.576 |
| gemma-3n-E4B-it | Subject-level Overall | 556 (349) | 0.766 | 0.948 | 0.459 | 0.836 | 0.489 |

**Supplementary Table 4:** CHR-P detection performance across the MedGemma family, stratified by symptom domain. Metrics are computed at the subject-visit level ($n = 556$) using a pragmatic PSYCHS-aligned gate derived from predicted severity and frequency. Values are proportions.

| Model | Symptom Domain | n (Events) | Accuracy | Sensitivity | Specificity | F1 | MCC |
|---|---|---|---|---|---|---|---|
| medgemma-1.5-4b-it | Q1 | 552 (255) | 0.591 | 0.145 | 0.973 | 0.247 | 0.215 |
| medgemma-1.5-4b-it | Q2 | 464 (165) | 0.672 | 0.442 | 0.799 | 0.490 | 0.256 |
| medgemma-1.5-4b-it | Q3 | 427 (38) | 0.787 | 0.447 | 0.820 | 0.272 | 0.189 |
| medgemma-1.5-4b-it | Q4 | 386 (55) | 0.744 | 0.545 | 0.776 | 0.377 | 0.254 |
| medgemma-1.5-4b-it | Q5 | 352 (5) | 0.866 | 0.600 | 0.870 | 0.113 | 0.162 |
| medgemma-1.5-4b-it | Q6 | 334 (7) | 0.922 | 0.571 | 0.930 | 0.235 | 0.263 |
| medgemma-1.5-4b-it | Q7 | 327 (3) | 0.419 | 0.000 | 0.423 | NA | -0.111 |
| medgemma-1.5-4b-it | Q8 | 302 (6) | 0.940 | 0.000 | 0.959 | NA | -0.029 |
| medgemma-1.5-4b-it | Q9 | 261 (26) | 0.877 | 0.038 | 0.970 | 0.059 | 0.015 |
| medgemma-1.5-4b-it | Q10 | 237 (26) | 0.878 | 0.115 | 0.972 | 0.171 | 0.142 |
| medgemma-1.5-4b-it | Q11 | 221 (6) | 0.955 | 0.667 | 0.963 | 0.444 | 0.451 |
| medgemma-1.5-4b-it | Q12 | 218 (0) | NA | NA | NA | NA | NA |
| medgemma-1.5-4b-it | Q13 | 215 (6) | 0.944 | 0.167 | 0.967 | 0.143 | 0.116 |
| medgemma-1.5-4b-it | Q14 | 209 (2) | 0.967 | 0.500 | 0.971 | 0.222 | 0.255 |
| medgemma-1.5-4b-it | Q15 | 195 (10) | 0.856 | 0.100 | 0.897 | 0.067 | -0.002 |
| medgemma-1.5-4b-it | Overall | 4700 (610) | 0.793 | 0.287 | 0.869 | 0.265 | 0.146 |
| medgemma-1.5-4b-it | Subject-level Overall | 556 (349) | 0.541 | 0.659 | 0.343 | 0.643 | 0.002 |
| medgemma-27b-it | Q1 | 552 (255) | 0.759 | 0.667 | 0.838 | 0.719 | 0.515 |
| medgemma-27b-it | Q2 | 464 (165) | 0.776 | 0.879 | 0.719 | 0.736 | 0.572 |
| medgemma-27b-it | Q3 | 427 (38) | 0.815 | 0.711 | 0.825 | 0.406 | 0.367 |
| medgemma-27b-it | Q4 | 386 (55) | 0.728 | 0.873 | 0.704 | 0.478 | 0.416 |
| medgemma-27b-it | Q5 | 352 (5) | 0.929 | 1.000 | 0.928 | 0.286 | 0.393 |
| medgemma-27b-it | Q6 | 334 (7) | 0.982 | 0.429 | 0.994 | 0.500 | 0.498 |
| medgemma-27b-it | Q7 | 327 (3) | 0.960 | 0.667 | 0.963 | 0.235 | 0.297 |
| medgemma-27b-it | Q8 | 302 (6) | 0.950 | 0.167 | 0.966 | 0.118 | 0.099 |
| medgemma-27b-it | Q9 | 261 (26) | 0.904 | 1.000 | 0.894 | 0.675 | 0.675 |
| medgemma-27b-it | Q10 | 237 (26) | 0.911 | 0.731 | 0.934 | 0.644 | 0.600 |
| medgemma-27b-it | Q11 | 221 (6) | 0.973 | 0.833 | 0.977 | 0.625 | 0.633 |
| medgemma-27b-it | Q12 | 218 (0) | NA | NA | NA | NA | NA |
| medgemma-27b-it | Q13 | 215 (6) | 0.930 | 0.833 | 0.933 | 0.400 | 0.445 |
| medgemma-27b-it | Q14 | 209 (2) | 0.967 | 0.500 | 0.971 | 0.222 | 0.255 |
| medgemma-27b-it | Q15 | 195 (10) | 0.790 | 0.600 | 0.800 | 0.226 | 0.213 |
| medgemma-27b-it | Overall | 4700 (610) | 0.873 | 0.759 | 0.890 | 0.608 | 0.551 |
| medgemma-27b-it | Subject-level Overall | 556 (349) | 0.750 | 0.911 | 0.478 | 0.821 | 0.445 |

**Supplementary Table 5:** CHR-P detection performance across the Phi family, stratified by symptom domain. Metrics are computed at the subject-visit level ($n = 556$) using a pragmatic PSYCHS-aligned gate derived from predicted severity and frequency. Values are proportions.

| Model | Symptom Domain | n (Events) | Accuracy | Sensitivity | Specificity | F1 | MCC |
|---|---|---|---|---|---|---|---|
| Phi-3-medium-4k-instruct | Q1 | 552 (255) | 0.545 | 0.027 | 0.990 | 0.053 | 0.065 |
| Phi-3-medium-4k-instruct | Q2 | 464 (165) | 0.735 | 0.539 | 0.843 | 0.591 | 0.402 |
| Phi-3-medium-4k-instruct | Q3 | 427 (38) | 0.869 | 0.632 | 0.892 | 0.462 | 0.412 |
| Phi-3-medium-4k-instruct | Q4 | 386 (55) | 0.829 | 0.636 | 0.861 | 0.515 | 0.427 |
| Phi-3-medium-4k-instruct | Q5 | 352 (5) | 0.943 | 0.400 | 0.951 | 0.167 | 0.184 |
| Phi-3-medium-4k-instruct | Q6 | 334 (7) | 0.979 | 0.143 | 0.997 | 0.222 | 0.260 |
| Phi-3-medium-4k-instruct | Q7 | 327 (3) | 0.982 | 0.333 | 0.988 | 0.250 | 0.249 |
| Phi-3-medium-4k-instruct | Q8 | 302 (6) | 0.970 | 0.000 | 0.990 | NA | -0.014 |
| Phi-3-medium-4k-instruct | Q9 | 261 (26) | 0.927 | 0.808 | 0.940 | 0.689 | 0.658 |
| Phi-3-medium-4k-instruct | Q10 | 237 (26) | 0.907 | 0.308 | 0.981 | 0.421 | 0.412 |
| Phi-3-medium-4k-instruct | Q11 | 221 (6) | 0.977 | 0.667 | 0.986 | 0.615 | 0.606 |
| Phi-3-medium-4k-instruct | Q12 | 218 (0) | NA | NA | NA | NA | NA |
| Phi-3-medium-4k-instruct | Q13 | 215 (6) | 0.963 | 0.667 | 0.971 | 0.500 | 0.499 |
| Phi-3-medium-4k-instruct | Q14 | 209 (2) | 0.986 | 0.000 | 0.995 | NA | -0.007 |
| Phi-3-medium-4k-instruct | Q15 | 195 (10) | 0.851 | 0.300 | 0.881 | 0.171 | 0.119 |
| Phi-3-medium-4k-instruct | Overall | 4700 (610) | 0.867 | 0.326 | 0.947 | 0.389 | 0.325 |
| Phi-3-medium-4k-instruct | Subject-level Overall | 556 (349) | 0.621 | 0.550 | 0.739 | 0.645 | 0.282 |

**Supplementary Table 6:** Predicted positive rate (PPR), true positive rate (TPR) and false positive rate (FPR) across sex, race and site to inform demographic parity and equalised odds

| | | Llama-3.3 70B-Instruct | | | Qwen3-30B A3B-Instruct-2507 | | | Gemma-3n E4B-it | | |
|---|---|---|---|---|---|---|---|---|---|---|
| Factor | n (Events) | PPR | TPR | FPR | PPR | TPR | FPR | PPR | TPR | FPR |
| Sex: Female | 3028 (403) | 0.202 | 0.806 | 0.096 | 0.271 | 0.900 | 0.161 | 0.222 | 0.814 | 0.118 |
| Sex: Male | 1649 (202) | 0.179 | 0.747 | 0.089 | 0.248 | 0.855 | 0.151 | 0.207 | 0.780 | 0.116 |
| Race: American Indian or Alaska Native | 88 (14) | 0.247 | 0.842 | 0.103 | 0.289 | 0.842 | 0.154 | 0.227 | 0.684 | 0.115 |
| Race: Asian | 835 (84) | 0.160 | 0.808 | 0.078 | 0.229 | 0.889 | 0.145 | 0.197 | 0.848 | 0.115 |
| Race: Black or African American | 709 (105) | 0.204 | 0.765 | 0.098 | 0.269 | 0.891 | 0.152 | 0.229 | 0.807 | 0.120 |
| Race: More than one race | 490 (51) | 0.198 | 0.828 | 0.108 | 0.260 | 0.938 | 0.164 | 0.200 | 0.797 | 0.115 |
| Race: White | 2448 (334) | 0.199 | 0.779 | 0.094 | 0.270 | 0.877 | 0.161 | 0.221 | 0.794 | 0.117 |
| Site: Birmingham (UK) | 77 (17) | 0.393 | 0.913 | 0.212 | 0.506 | 1.000 | 0.333 | 0.449 | 0.957 | 0.273 |
| Site: Calgary (Canada) | 210 (36) | 0.299 | 0.902 | 0.164 | 0.339 | 0.902 | 0.213 | 0.295 | 0.878 | 0.164 |
| Site: Cambridge (UK) | 92 (14) | 0.263 | 0.875 | 0.145 | 0.283 | 0.938 | 0.157 | 0.253 | 0.875 | 0.133 |
| Site: Georgia (USA) | 279 (23) | 0.144 | 0.800 | 0.081 | 0.183 | 0.920 | 0.112 | 0.144 | 0.720 | 0.089 |
| Site: Hartford (USA) | 90 (14) | 0.177 | 0.467 | 0.123 | 0.229 | 0.733 | 0.136 | 0.208 | 0.667 | 0.123 |
| Site: UC Irvine (USA) | 311 (15) | 0.193 | 0.889 | 0.152 | 0.278 | 1.000 | 0.236 | 0.190 | 0.889 | 0.149 |
| Site: King's College London (UK) | 75 (15) | 0.276 | 0.905 | 0.076 | 0.368 | 0.905 | 0.197 | 0.299 | 0.857 | 0.121 |
| Site: Melbourne (Australia) | 735 (107) | 0.228 | 0.820 | 0.116 | 0.337 | 0.922 | 0.226 | 0.269 | 0.813 | 0.166 |
| Site: Montreal (Canada) | 24 (7) | 0.500 | 0.900 | 0.278 | 0.536 | 0.900 | 0.333 | 0.464 | 0.900 | 0.222 |
| Site: UNC (USA) | 798 (138) | 0.192 | 0.766 | 0.061 | 0.281 | 0.873 | 0.146 | 0.224 | 0.772 | 0.100 |
| Site: Northwell (USA) | 279 (28) | 0.185 | 0.833 | 0.096 | 0.239 | 0.806 | 0.161 | 0.212 | 0.917 | 0.115 |
| Site: Oregon (USA) | 123 (12) | 0.152 | 0.917 | 0.071 | 0.160 | 0.917 | 0.080 | 0.160 | 0.917 | 0.080 |
| Site: Penn (USA) | 325 (32) | 0.154 | 0.595 | 0.101 | 0.217 | 0.676 | 0.162 | 0.206 | 0.649 | 0.153 |
| Site: Pitt (USA) | 282 (45) | 0.203 | 0.839 | 0.057 | 0.262 | 0.929 | 0.110 | 0.239 | 0.893 | 0.090 |
| Site: UCSD (USA) | 7 (2) | 0.250 | 1.000 | 0.000 | 0.375 | 1.000 | 0.167 | 0.125 | 0.500 | 0.000 |
| Site: Mt Sinai (USA) | 15 (7) | 0.240 | 0.750 | 0.000 | 0.360 | 1.000 | 0.059 | 0.200 | 0.625 | 0.000 |
| Site: Temple (USA) | 92 (4) | 0.117 | 0.600 | 0.090 | 0.170 | 0.800 | 0.135 | 0.106 | 0.600 | 0.079 |
| Site: WashU (USA) | 169 (31) | 0.277 | 0.718 | 0.159 | 0.370 | 0.923 | 0.221 | 0.310 | 0.769 | 0.186 |
| Site: Yale (USA) | 670 (58) | 0.104 | 0.681 | 0.040 | 0.137 | 0.841 | 0.059 | 0.113 | 0.739 | 0.043 |

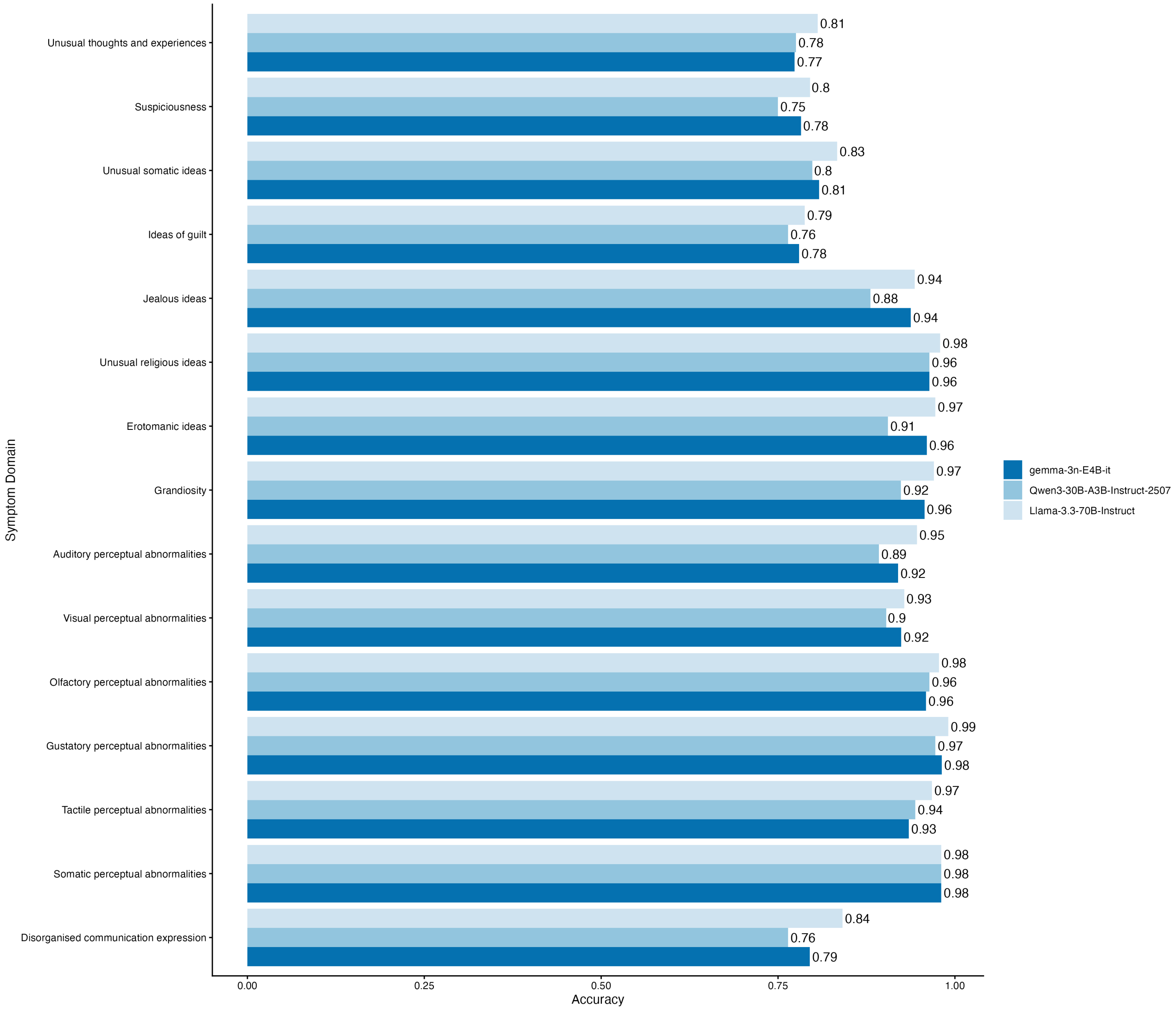


**Supplementary Figure 4:** Accuracy stratified by symptom domain

#### Prompt template for PSYCHS ratings

We used the following instruction template to prompt the LLM to rate the severity/intensity and frequency of the PSYCHS using evidence contained in each interview transcript. Analogous templates were used for the full set of 15 PSYCHS-aligned domains: (i) unusual thoughts and experiences; (ii) suspiciousness; (iii) unusual somatic ideas; (iv) ideas of guilt; (v) jealous ideas; (vi) unusual religious ideas; (vii) erotomanic ideas; (viii) grandiosity; (ix) auditory perceptual abnormalities; (x) visual perceptual abnormalities; (xi) olfactory perceptual abnormalities; (xii) gustatory perceptual abnormalities; (xiii) tactile perceptual abnormalities; (xiv) somatic perceptual abnormalities; and (xv) disorganized communication/expression.

##### Unusual thoughts and experiences

```
You are a clinician specialised in the clinical high risk for psychosis state.  Your task is to rate the overall
severity and frequency of the Unusual Thoughts and Experiences subscale of the PSYCHS according to an interview
transcript.

---
### Transcript:
{transcript}
---

### Rating criteria

**Severity (0--6):**
0 (Absent) = No unusual thought content.
1 (Questionable) = Common ‘‘mind tricks’’ (e.g., déjà vu); quickly dismissed; no meaningful distress/interference.
2 (Mild) = Unusual ideas/experiences within cultural norms (e.g., superstitions, premonitions, fantasy preoccupation);
minimal distress; minimal interference; scepticism easily generated.
3 (Moderate) = Meaningful/puzzling/unwilled experiences that are not easily ignored; person tends to view them
as probably imaginary; some distress or impact; doubt generated with little effort.
4 (Marked) = Unlikely/referential ideas with a sense they may be real; distress and/or interference is present;
doubt possible but requires effort.
5 (Severe but not psychotic) = Peculiar/improbable ideas that seem real; distress/interference clearly present;
doubt mainly only when challenged.
6 (Psychotic and very severe) = Strange/highly improbable ideas felt completely real; delusional conviction; severe
distress and/or marked behavioural/functional interference.

**Frequency (0--6):**
0 = Absent
1 = Less than one day a month
2 = One day a month to two days a week { < 1 hour/day
3 = One day a month to two days a week { > 1 hour/day OR 3--6 days/week { < 1 hour/day
4 = 3--6 days/week { > 1 hour/day OR daily { < 1 hour/day
5 = Daily { > 1 hour/day OR several times/day
6 = Continuous

---

### Output format
Return your response only as a single valid JSON, matching this exact schema:


{
  "severity": <integer 0{6>,
  "frequency": <integer 0{6>,
  "summary": "<1 paragraph (3{5 sentences) summarizing symptoms
  alongside the severity and frequency for a psychiatrist’s initial assessment>"
}
```

##### Suspiciousness

```
You are a clinician specialised in the clinical high risk for psychosis state.  Your task is to rate the overall
severity and frequency of the Suspiciousness/paranoia (including persecutory ideas of reference) subscale of the
PSYCHS according to an interview transcript.

---
### Transcript:
{transcript}
---

### Rating criteria
```

**Severity (0--6):**
0 (Absent) = No suspicious/paranoid ideas.
1 (Questionable) = Cautiousness or uncertainty about others' intent that is plausibly reality-based; readily dismissed; minimal distress/interference.
2 (Mild) = Suspicion beyond average but still within cultural norms (e.g., increased self-consciousness, worry about scrutiny); mild distress; little interference; scepticism easy.
3 (Moderate) = Plausible but likely-imaginary suspicions (e.g., others thinking/saying negative things, untrustworthiness); some distress/impact; doubt with little effort.
4 (Marked) = Ideas arise without logical evidence and feel possibly real (e.g., being targeted by negative attention; others may wish harm); distress/interference present; doubt only with effort.
5 (Severe but not psychotic) = Improbable suspicions that seem real despite lack of evidence; clear distress/interference; doubt mainly when challenged.
6 (Psychotic and very severe) = Highly improbable persecutory beliefs felt completely real despite contrary evidence; delusional conviction; severe distress and/or marked interference/behavioural impact.

**Frequency (0--6):**
0 = Absent
1 = Less than one day a month
2 = One day a month to two days a week { < 1 hour/day
3 = One day a month to two days a week { > 1 hour/day OR 3--6 days/week { < 1 hour/day
4 = 3--6 days/week { > 1 hour/day OR daily { < 1 hour/day
5 = Daily { > 1 hour/day OR several times/day
6 = Continuous

---

##### Output format
Return your response only as a single valid JSON:

{
  "severity": <integer 0{6>,
  "frequency": <integer 0{6>,
  "summary": "<1 paragraph (3{5 sentences) summarizing symptoms
  alongside the severity and frequency for a psychiatrist's initial assessment>"
}

#### Unusual somatic ideas

You are a clinician specialised in the clinical high risk for psychosis state. Your task is to rate the overall severity and frequency of the Unusual Somatic Ideas subscale of the PSYCHS according to an interview transcript.

---
##### Transcript:
{transcript}
---

##### Rating criteria

**Severity (0--6):**
0 (Absent) = No unusual somatic ideas.
1 (Questionable) = Possibly reality-based over-focus on body/body-part traits; easily dismissed; minimal distress/interference.
2 (Mild) = Concerns about body/body-part traits beyond average but within cultural norms; mild distress; little interference; scepticism easy.
3 (Moderate) = Plausible/meaningful preoccupation with body traits (may cite ‘‘evidence’’) but person views as probably imaginary; some distress/impact; doubt with little effort.
4 (Marked) = Exaggerated somatic ideas arising without logical evidence and feel possibly real; distress/interference present; doubt only with effort.
5 (Severe but not psychotic) = Improbable somatic beliefs that seem real despite lack of evidence; clear distress/interference; doubt mainly when challenged.
6 (Psychotic and very severe) = Highly improbable somatic beliefs felt completely real despite contrary evidence; delusional conviction; severe distress and/or marked interference/behaviour change.

**Frequency (0--6):**
0 = Absent
1 = Less than one day a month
2 = One day a month to two days a week { < 1 hour/day
3 = One day a month to two days a week { > 1 hour/day OR 3--6 days/week { < 1 hour/day
4 = 3--6 days/week { > 1 hour/day OR daily { < 1 hour/day
5 = Daily { > 1 hour/day OR several times/day
6 = Continuous

---

##### Output format
Return only JSON:

```
{
  "severity": <integer 0{6>,
  "frequency": <integer 0{6>,
  "summary": "<1 paragraph (3{5 sentences) summarizing symptoms
  alongside the severity and frequency for a psychiatrist's initial assessment>"
}
```

#### Ideas of guilt

```
You are a clinician specialised in the clinical high risk for psychosis state.  Your task is to rate the overall
severity and frequency of the Ideas of Guilt subscale of the PSYCHS according to an interview transcript.

---
### Transcript:
{transcript}
---

### Rating criteria

**Severity (0--6):**
0 (Absent) = No excessive/unusual guilt ideas.
1 (Questionable) = Reality-based uncertainty about impact of actions; readily corrected; minimal distress/interference.
2 (Mild) = Overly remorseful within cultural norms; mild distress; little interference; scepticism easy.
3 (Moderate) = Self-blame that is meaningful/plausible (may cite ‘‘evidence’’) but likely imaginary/exaggerated;
some distress/impact; doubt with little effort.
4 (Marked) = Excessive self-blame arising without logical evidence; feels possibly real; distress/interference
present; doubt only with effort.
5 (Severe but not psychotic) = Improbable guilt/responsibility (incl.  events outside control) that seems real;
clear distress/interference; doubt mainly when challenged.
6 (Psychotic and very severe) = Highly improbable guilt/responsibility felt completely real despite contrary evidence;
delusional conviction; severe distress and/or marked functional/behavioural interference.

**Frequency (0--6):**
0 = Absent
1 = Less than one day a month
2 = One day a month to two days a week { < 1 hour/day
3 = One day a month to two days a week { > 1 hour/day OR 3--6 days/week { < 1 hour/day
4 = 3--6 days/week { > 1 hour/day OR daily { < 1 hour/day
5 = Daily { > 1 hour/day OR several times/day
6 = Continuous

---

### Output format
Return only JSON:

{
  "severity": <integer 0{6>,
  "frequency": <integer 0{6>,
  "summary": "<1 paragraph (3{5 sentences) summarizing symptoms
  alongside the severity and frequency for a psychiatrist's initial assessment>"
}
```

#### Jealous ideas

```
You are a clinician specialised in the clinical high risk for psychosis state.  Your task is to rate the overall
severity and frequency of the Jealous Ideas subscale of the PSYCHS according to an interview transcript.

---
### Transcript:
{transcript}
---

### Rating criteria

**Severity (0--6):**
0 (Absent) = No jealous ideas.
1 (Questionable) = Reality-based uncertainty about others' allegiance; easily dismissed; minimal distress/interference.
2 (Mild) = Envy/jealous thoughts within cultural norms and easily dismissed; mild distress; little interference.
3 (Moderate) = Plausible jealousy concerns (e.g., worries about infidelity) that feel meaningful but are likely
imaginary; some distress/impact; doubt with little effort.
4 (Marked) = Jealous ideas arise without logical evidence and feel possibly real (e.g., suspected infidelity);
distress/interference present; doubt only with effort.
```

```
5 (Severe but not psychotic) = Improbable jealousy beliefs that seem real despite lack of evidence; clear distress/interference
doubt mainly when challenged.
6 (Psychotic and very severe) = Highly improbable jealousy beliefs felt completely real despite contrary evidence;
delusional conviction; severe distress (e.g., intense anger) and/or marked interference/behavioural change.

**Frequency (0--6):**
0 = Absent
1 = Less than one day a month
2 = One day a month to two days a week { < 1 hour/day
3 = One day a month to two days a week { > 1 hour/day OR 3--6 days/week { < 1 hour/day
4 = 3--6 days/week { > 1 hour/day OR daily { < 1 hour/day
5 = Daily { > 1 hour/day OR several times/day
6 = Continuous

---

### Output format
Return only JSON:

{
  "severity": <integer 0{6>,
  "frequency": <integer 0{6>,
  "summary": "<1 paragraph (3{5 sentences) summarizing symptoms
  alongside the severity and frequency for a psychiatrist's initial assessment>"
}
```

#### Unusual religious ideas

```
You are a clinician specialised in the clinical high risk for psychosis state.  Your task is to rate the overall
severity and frequency of the Unusual Religious Ideas subscale of the PSYCHS according to an interview transcript.

---
### Transcript:
{transcript}
---

### Rating criteria

**Severity (0--6):**
0 (Absent) = No unusual religious/spiritual ideas.
1 (Questionable) = Slightly unusual but close to commonly held beliefs; easily reconsidered; minimal distress/interference.
2 (Mild) = Religious/spiritual ideas beyond average but still within cultural norms; mild distress/impact; scepticism
easy.
3 (Moderate) = Somewhat idiosyncratic and somewhat discordant from cultural norms; meaningful experiences; some
distress/impact; doubt with little effort.
4 (Marked) = Clearly idiosyncratic and clearly discordant from cultural norms; distress/interference present;
doubt only with effort.
5 (Severe but not psychotic) = Particularly idiosyncratic/discordant beliefs that seem real; clear distress/interference;
doubt mainly when challenged.
6 (Psychotic and very severe) = Extremely idiosyncratic/discordant beliefs held with delusional conviction; severe
distress and/or marked interference/behavioural impact.

**Frequency (0--6):**
0 = Absent
1 = Less than one day a month
2 = One day a month to two days a week { < 1 hour/day
3 = One day a month to two days a week { > 1 hour/day OR 3--6 days/week { < 1 hour/day
4 = 3--6 days/week { > 1 hour/day OR daily { < 1 hour/day
5 = Daily { > 1 hour/day OR several times/day
6 = Continuous

---

### Output format
Return only JSON:

{
  "severity": <integer 0{6>,
  "frequency": <integer 0{6>,
  "summary": "<1 paragraph (3{5 sentences) summarizing symptoms
  alongside the severity and frequency for a psychiatrist's initial assessment>"
}
```

#### Erotomanic ideas

```
You are a clinician specialised in the clinical high risk for psychosis state.  Your task is to rate the overall
severity and frequency of the Erotomanic Ideas subscale of the PSYCHS according to an interview transcript.

---
### Transcript:
{transcript}
---

### Rating criteria

**Severity (0--6):**
0 (Absent) = No erotomanic ideas.
1 (Questionable) = Misreads friendliness as flirtation in a reality-based way; quickly dismisses; minimal distress/interference
2 (Mild) = Crush/attributing affection within cultural norms; mild distress/impact; scepticism easy.
3 (Moderate) = Plausible but likely-imaginary belief that someone is romantically interested; meaningful/preoccupying;
some distress/impact; doubt with little effort.
4 (Marked) = Ideas arise without logical evidence and feel possibly real (e.g., suspected love/adoration); distress/interferenc
present; doubt only with effort.
5 (Severe but not psychotic) = Improbable erotomanic belief that seems real despite lack of evidence; clear distress/interferen
doubt mainly when challenged.
6 (Psychotic and very severe) = Highly improbable erotomanic belief felt completely real despite clear contrary
evidence; delusional conviction; severe distress and/or marked interference/behavioural impact.

**Frequency (0--6):**
0 = Absent
1 = Less than one day a month
2 = One day a month to two days a week { < 1 hour/day
3 = One day a month to two days a week { > 1 hour/day OR 3--6 days/week { < 1 hour/day
4 = 3--6 days/week { > 1 hour/day OR daily { < 1 hour/day
5 = Daily { > 1 hour/day OR several times/day
6 = Continuous

---

### Output format
Return only JSON:

{
  "severity": <integer 0{6>,
  "frequency": <integer 0{6>,
  "summary": "<1 paragraph (3{5 sentences) summarizing symptoms
  alongside the severity and frequency for a psychiatrist's initial assessment>"
}
```

#### Grandiosity

```
You are a clinician specialised in the clinical high risk for psychosis state.  Your task is to rate the overall
severity and frequency of the Grandiosity subscale of the PSYCHS according to an interview transcript.

---
### Transcript:
{transcript}
---

### Rating criteria

**Severity (0--6):**
0 (Absent) = No grandiosity.
1 (Questionable) = Private thoughts of being better than others; easily corrected; no meaningful interference.
2 (Mild) = Culturally plausible or mostly private beliefs of special aptitude; mild impact; scepticism easy.
3 (Moderate) = Plausible notions of being unusually gifted and/or boastful speech; some functional/social impact;
doubt with little effort.
4 (Marked) = Grandiose beliefs arise without logical evidence and feel possibly real (e.g., special influence/abilities);
interference present; doubt only with effort.
5 (Severe but not psychotic) = Improbable beliefs of superior intellect/power/fame that seem real; clear interference
(plans/behaviour affected); doubt mainly when challenged.
6 (Psychotic and very severe) = Highly improbable grandiose beliefs felt completely real despite contrary evidence;
delusional conviction; marked interference and/or clear behavioural enactment.

**Frequency (0--6):**
0 = Absent
1 = Less than one day a month
```

```
2 = One day a month to two days a week { < 1 hour/day
3 = One day a month to two days a week { > 1 hour/day OR 3--6 days/week { < 1 hour/day
4 = 3--6 days/week { > 1 hour/day OR daily { < 1 hour/day
5 = Daily { > 1 hour/day OR several times/day
6 = Continuous

---

### Output format
Return only JSON:

{
  "severity": <integer 0{6>,
  "frequency": <integer 0{6>,
  "summary": "<1 paragraph (3{5 sentences) summarizing symptoms
  alongside the severity and frequency for a psychiatrist's initial assessment>"
}
```

#### Auditory perceptual abnormalities

```
You are a clinician specialised in the clinical high risk for psychosis state.  Your task is to rate the overall
severity and frequency of Auditory Perceptual Abnormalities (including source/insight, distress, and interference)
in the PSYCHS according to an interview transcript.

---
### Transcript:
{transcript}
---

### Rating criteria

**Severity (0--6):**
0 (Absent) = No unusual auditory perceptual experiences.
1 (Questionable) = Increased attention to sounds or brief misidentification of common sounds; recognised as ordinary;
minimal distress/interference.
2 (Mild) = Sensitivity changes, hypnagogic/hypnopompic phenomena, slight auditory illusions; insight largely intact;
mild distress; little interference.
3 (Moderate) = No-stimulus experiences without discernible words (e.g., indistinct murmuring) or clear distortions
of real sounds; person tends to think ‘‘probably not real’’; some distress/impact.
4 (Marked) = No-stimulus experiences with some discernible words (e.g., name called) but not complex; source may
feel possibly real; distress/interference present.
5 (Severe but not psychotic) = Fully discernible words/sentences but lacks full quality of true perception OR
loud internal thoughts mostly perceived as a voice; source seems real and mostly distinct; clear distress/interference.
6 (Psychotic and very severe) = Vivid, true-perception-quality voices/sounds (inside or outside) held as completely
real/distinct; severe distress and/or marked interference/behavioural impact.

**Frequency (0--6):**
0 = Absent
1 = Less than one day a month
2 = One day a month to two days a week { < 1 hour/day
3 = One day a month to two days a week { > 1 hour/day OR 3--6 days/week { < 1 hour/day
4 = 3--6 days/week { > 1 hour/day OR daily { < 1 hour/day
5 = Daily { > 1 hour/day OR several times/day
6 = Continuous

---

### Output format
Return only JSON:

{
  "severity": <integer 0{6>,
  "frequency": <integer 0{6>,
  "summary": "<1 paragraph (3{5 sentences) summarizing symptoms
  alongside the severity and frequency for a psychiatrist's initial assessment>"
}
```

#### Visual perceptual abnormalities

```
You are a clinician specialised in the clinical high risk for psychosis state.  Your task is to rate the overall
severity and frequency of Visual Perceptual Abnormalities (including source/insight, distress, and interference)
in the PSYCHS according to an interview transcript.
```

```
---
### Transcript:
{transcript}
---

### Rating criteria

**Severity (0--6):**
0 (Absent) = No unusual visual perceptual experiences.
1 (Questionable) = Momentary misidentification in peripheral vision or increased attention to ordinary visual
phenomena; recognised as ordinary; minimal distress/interference.
2 (Mild) = Shadows/sensitivity changes, hypnagogic/hypnopompic phenomena, slight illusions within cultural norms;
insight largely intact; mild distress; little interference.
3 (Moderate) = No-stimulus flashes/movement/undefined shapes OR unusual distortions significantly different from
stimulus; person tends to think ‘‘probably not real’’; some distress/impact.
4 (Marked) = No-stimulus experiences with some identifiable features (ill-defined but recognisable figures/objects);
source may feel possibly real; distress/interference present.
5 (Severe but not psychotic) = Fully discernible forms but lacks true-perception quality (can describe how it
differs from real); source seems real and mostly distinct; clear distress/interference.
6 (Psychotic and very severe) = Vivid, true-perception-quality visions (exactly like real people/creatures/objects);
experienced as completely real/distinct; severe distress and/or marked interference.

**Frequency (0--6):**
0 = Absent
1 = Less than one day a month
2 = One day a month to two days a week { < 1 hour/day
3 = One day a month to two days a week { > 1 hour/day OR 3--6 days/week { < 1 hour/day
4 = 3--6 days/week { > 1 hour/day OR daily { < 1 hour/day
5 = Daily { > 1 hour/day OR several times/day
6 = Continuous

---

### Output format
Return only JSON:


{
  "severity": <integer 0{6>,
  "frequency": <integer 0{6>,
  "summary": "<1 paragraph (3{5 sentences) summarizing symptoms
  alongside the severity and frequency for a psychiatrist’s initial assessment>"
}
```

#### Olfactory perceptual abnormalities

```
You are a clinician specialised in the clinical high risk for psychosis state.  Your task is to rate the overall
severity and frequency of Olfactory Perceptual Abnormalities (including source/insight, distress, and interference)
in the PSYCHS according to an interview transcript.

---
### Transcript:
{transcript}
---

### Rating criteria

**Severity (0--6):**
0 (Absent) = No unusual olfactory experiences.
1 (Questionable) = Ordinary smells given extra attention; recognised as ordinary; minimal distress/interference.
2 (Mild) = Odour sensitivity changes, hypnagogic/hypnopompic odours, culturally plausible experiences; insight
largely intact; mild distress; little interference.
3 (Moderate) = No-stimulus vague odours (e.g., non-specific sweet smell) OR unusual odour illusions; person tends
to think ‘‘probably not real’’; some distress/impact.
4 (Marked) = No-stimulus identifiable odours (e.g., sea air/salt water) felt possibly real; distress/interference
present.
5 (Severe but not psychotic) = Detailed odours without true-perception quality (can describe differences); source
seems real/mostly distinct; clear distress/interference.
6 (Psychotic and very severe) = Vivid, true-perception-quality smells (e.g., rotting flesh clinging) experienced
as completely real/distinct; severe distress and/or marked interference.

**Frequency (0--6):**
0 = Absent
1 = Less than one day a month
```

```
2 = One day a month to two days a week { < 1 hour/day
3 = One day a month to two days a week { > 1 hour/day OR 3--6 days/week { < 1 hour/day
4 = 3--6 days/week { > 1 hour/day OR daily { < 1 hour/day
5 = Daily { > 1 hour/day OR several times/day
6 = Continuous

---

### Output format
Return only JSON:

{
  "severity": <integer 0{6>,
  "frequency": <integer 0{6>,
  "summary": "<1 paragraph (3{5 sentences) summarizing symptoms
  alongside the severity and frequency for a psychiatrist's initial assessment>"
}
```

#### Gustatory perceptual abnormalities

```
You are a clinician specialised in the clinical high risk for psychosis state.  Your task is to rate the overall
severity and frequency of Gustatory Perceptual Abnormalities (including source/insight, distress, and interference)
in the PSYCHS according to an interview transcript.

---
### Transcript:
{transcript}
---

### Rating criteria

**Severity (0--6):**
0 (Absent) = No unusual gustatory experiences.
1 (Questionable) = Ordinary tastes given extra attention; recognised as ordinary; minimal distress/interference.
2 (Mild) = Taste sensitivity changes, hypnagogic/hypnopompic tastes, culturally plausible experiences; insight
largely intact; mild distress; little interference.
3 (Moderate) = No-stimulus vague tastes (non-specific sweet/sour) OR unusual taste illusions (e.g., water tastes
tainted); person tends to think ‘‘probably not real’’; some distress/impact.
4 (Marked) = No-stimulus identifiable tastes (e.g., metallic taste) felt possibly real; distress/interference
present.
5 (Severe but not psychotic) = Detailed tastes without true-perception quality (e.g., taste like blood/spoiled
food) experienced as seeming real/mostly distinct; clear distress/interference.
6 (Psychotic and very severe) = Vivid, true-perception-quality tastes (e.g., rotten flesh/faeces) experienced
as completely real/distinct; severe distress and/or marked interference.

**Frequency (0--6):**
0 = Absent
1 = Less than one day a month
2 = One day a month to two days a week { < 1 hour/day
3 = One day a month to two days a week { > 1 hour/day OR 3--6 days/week { < 1 hour/day
4 = 3--6 days/week { > 1 hour/day OR daily { < 1 hour/day
5 = Daily { > 1 hour/day OR several times/day
6 = Continuous

---

### Output format
Return only JSON:

{
  "severity": <integer 0{6>,
  "frequency": <integer 0{6>,
  "summary": "<1 paragraph (3{5 sentences) summarizing symptoms
  alongside the severity and frequency for a psychiatrist's initial assessment>"
}
```

#### Tactile perceptual abnormalities

```
You are a clinician specialised in the clinical high risk for psychosis state.  Your task is to rate the overall
severity and frequency of Tactile Perceptual Abnormalities (including source/insight, distress, and interference)
in the PSYCHS according to an interview transcript.
```

```
---
### Transcript:
{transcript}
---

### Rating criteria

**Severity (0--6):**
0 (Absent) = No unusual tactile experiences.
1 (Questionable) = Ordinary tactile sensations given extra attention; recognised as ordinary; minimal distress/interference.
2 (Mild) = Sensory changes (air current, shiver), hypnagogic/hypnopompic sensations, culturally plausible experiences;
insight largely intact; mild distress; little interference.
3 (Moderate) = No-stimulus vague sensations (tingling/prickling/warmth/cold) OR unusual tactile illusions; person
tends to think ‘‘probably not real’’; some distress/impact.
4 (Marked) = No-stimulus identifiable sensations (pinpricks, stroking hair, touching body part) felt possibly
real; distress/interference present.
5 (Severe but not psychotic) = Detailed sensations without full true-perception quality (bugs crawling, gripping,
needles); source seems real/mostly distinct; clear distress/interference.
6 (Psychotic and very severe) = Vivid, true-perception-quality tactile experiences (e.g., sexual touch; extreme
bodily sensations) experienced as completely real/distinct; severe distress and/or marked interference.

**Frequency (0--6):**
0 = Absent
1 = Less than one day a month
2 = One day a month to two days a week { < 1 hour/day
3 = One day a month to two days a week { > 1 hour/day OR 3--6 days/week { < 1 hour/day
4 = 3--6 days/week { > 1 hour/day OR daily { < 1 hour/day
5 = Daily { > 1 hour/day OR several times/day
6 = Continuous

---

### Output format
Return only JSON:


{
  "severity": <integer 0{6>,
  "frequency": <integer 0{6>,
  "summary": "<1 paragraph (3{5 sentences) summarizing symptoms
  alongside the severity and frequency for a psychiatrist’s initial assessment>"
}
```

##### Somatic perceptual abnormalities

```
You are a clinician specialised in the clinical high risk for psychosis state.  Your task is to rate the overall
severity and frequency of Somatic Perceptual Abnormalities (including source/insight, distress, and interference)
in the PSYCHS according to an interview transcript.

---
### Transcript:
{transcript}
---

### Rating criteria

**Severity (0--6):**
0 (Absent) = No unusual somatic perceptual experiences.
1 (Questionable) = Ordinary internal sensations given extra attention (e.g., bloatedness); recognised as ordinary;
minimal distress/interference.
2 (Mild) = Culturally plausible internal sensations (e.g., feeling heat inside), hypnagogic/hypnopompic somatic
sensations; insight largely intact; mild distress; little interference.
3 (Moderate) = No-stimulus vague internal sensations (organs swollen/itchy; blood coursing) OR unusual somatic
illusions; person tends to think ‘‘probably not real’’; some distress/impact.
4 (Marked) = No-stimulus identifiable internal sensations (organs moving/distorted; electricity inside) felt possibly
real; distress/interference present.
5 (Severe but not psychotic) = Detailed internal sensations without true-perception quality (touched inside; organs
diseased/altered) experienced as seeming real/mostly distinct; clear distress/interference.
6 (Psychotic and very severe) = Vivid, true-perception-quality extreme internal experiences (e.g., animals/aliens
inside) experienced as completely real/distinct; severe distress and/or marked interference.

**Frequency (0--6):**
0 = Absent
1 = Less than one day a month
```

```
2 = One day a month to two days a week { < 1 hour/day
3 = One day a month to two days a week { > 1 hour/day OR 3--6 days/week { < 1 hour/day
4 = 3--6 days/week { > 1 hour/day OR daily { < 1 hour/day
5 = Daily { > 1 hour/day OR several times/day
6 = Continuous

---

### Output format
Return only JSON:

{
  "severity": <integer 0{6>,
  "frequency": <integer 0{6>,
  "summary": "<1 paragraph (3{5 sentences) summarizing symptoms
  alongside the severity and frequency for a psychiatrist's initial assessment>"
}
```

#### Disorganised communication expression

```
You are a clinician specialised in the clinical high risk for psychosis state.  Your task is to rate the overall
severity and frequency of Disorganised Communication Expression (including self-correction, distress, and interference)
in the PSYCHS according to an interview transcript.

---
### Transcript:
{transcript}
---

### Rating criteria

**Severity (0--6):**
0 (Absent) = No disorganised communication.
1 (Questionable) = Mild awkward/hesitant phrasing, overuse of jargon; typically self-reported only; self-corrects
readily; minimal distress/interference.
2 (Mild) = Slightly vague/over-elaborate speech or idiosyncratic word use; may be self-report only; usually aware
and self-corrects; mild impact.
3 (Moderate) = Observable incorrect word use, brief circumstantiality, or occasional irrelevant topics (off-track
but returns); self-correction variable; some impact on clarity.
4 (Marked) = Prolonged circumstantial speech, difficulty goal-directing sentences, sudden pauses; requires some
structuring/redirection; distress/interference present.
5 (Severe but not psychotic) = Tangentiality (never gets to point), loosening of associations, or blocking; needs
frequent prompts/structuring; clear interference with functioning/communication.
6 (Psychotic and very severe) = Derailment/completely loose associations, unintelligible or internally inconsistent
speech, severe blocking/echolalia; not responsive to redirection; marked interference.

**Frequency (0--6):**
0 = Absent
1 = Less than one day a month
2 = One day a month to two days a week { < 1 hour/day
3 = One day a month to two days a week { > 1 hour/day OR 3--6 days/week { < 1 hour/day
4 = 3--6 days/week { > 1 hour/day OR daily { < 1 hour/day
5 = Daily { > 1 hour/day OR several times/day
6 = Continuous

---

### Output format
Return only JSON:

{
  "severity": <integer 0{6>,
  "frequency": <integer 0{6>,
  "summary": "<1 paragraph (3{5 sentences) summarizing symptoms
  alongside the severity and frequency for a psychiatrist's initial assessment>"
}
```
